# The Association of Social Distancing, Population Density, and Temperature with the SARS-CoV-2 Instantaneous Reproduction Number in Counties Across the United States

**DOI:** 10.1101/2020.05.08.20094474

**Authors:** David Rubin, Jing Huang, Brian T. Fisher, Antonio Gasparrini, Vicky Tam, Lihai Song, Xi Wang, Jason Kaufman, Kate Fitzpatrick, Arushi Jain, Heather Griffis, Koby Crammer, Gregory Tasian

**Affiliations:** Department of Pediatrics, Children’s Hospital of Philadelphia, Philadelphia, PA; Department of Pediatrics, Perelman School of Medicine at the University of Pennsylvania, Philadelphia, PA; Department of Biostatistics, Epidemiology, and Informatics, Perelman School of Medicine at the University of Pennsylvania, Philadelphia, PA; Department of Pediatrics, Division of Infectious Disease, Perelman School of Medicine at the University of Pennsylvania, Philadelphia, PA; Department of Public Health Environments and Society, London School of Hygiene and Tropical Medicine, London, UK; Department of Biomedical and Health Informatics, Data Science and Biostatistics Unit, Children’s Hospital of Philadelphia, Philadelphia, PA; Department of Surgery, Division of Urology, Children’s Hospital of Philadelphia, Philadelphia, PA; Department of Electrical Engineering, The Technion, Haifa, Israel; Centre for Statistical Methodology, London School of Hygiene and Tropical Medicine, London, UK; Centre on Climate Change and Planetary Health, London School of Hygiene and Tropical Medicine, London, UK; Center for Pediatric Clinical Effectiveness, Children’s Hospital of Philadelphia, Philadelphia, PA; PolicyLab, Children’s Hospital of Philadelphia, Philadelphia, PA; Perelman School of Medicine at the University of Pennsylvania, Philadelphia, PA

## Abstract

**Importance:** The Covid-19 pandemic has been marked by considerable heterogeneity in outbreaks across the United States. Local factors that may be associated with variation in SARS-CoV-2 transmission have not been well studied.

**Objective:** To examine the association of county-level factors with variation in the SARS-CoV-2 reproduction number over time.

**Design:** Observational study

**Setting:** 211 counties in 46 states and the District of Columbia between February 25, 2020 and April 23, 2020.

**Participants:** Residents within the counties (55% of the US population)

**Exposures:** Social distancing as measured by percent change in visits to non-essential businesses, population density, lagged daily wet bulb temperatures.

**Main Outcomes and Measures:** The instantaneous reproduction number (R_t_) which is the estimated number of cases generated by one case at a given time during the pandemic.

**Results:** Median case incidence was 1185 cases and fatality rate was 43.7 deaths per 100,000 people for the top decile of 21 counties, nearly ten times the incidence and fatality rate in the lowest density quartile. Average R_t_ in the first two weeks was 5.7 (SD 2.5) in the top decile, compared to 3.1 (SD 1.2) in the lowest quartile. In multivariable analysis, a 50% decrease in visits to non-essential businesses was associated with a 57% decrease in R_t_ (95% confidence interval, 56% to 58%). Cumulative temperature effects over 4 to 10 days prior to case incidence were nonlinear; relative R_t_ decreased as temperatures warmed above 32°F to 53°F, which was the point of minimum R_t_, then increased between 53°F and 66°F, at which point R_t_ began to decrease. At 55°F, and with a 70% reduction in visits to non-essential business, 96% of counties were estimated to fall below a threshold R_t_ of 1.0, including 86% of counties among the top density decile and 98% of counties in the lowest density quartile.

**Conclusions and Relevance:** Social distancing, lower population density, and temperate weather change were associated with a decreased SARS-Co-V-2 R_t_ in counties across the United States. These relationships can inform selective public policy planning in communities during the SARS-CoV-2 pandemic.

**Key Points:** *Question:* How is the instantaneous reproduction number (R_t_) of SARS-CoV-2 influenced by local area effects of social distancing, wet bulb temperature, and population density in counties across the United States?

*Findings:* Social distancing, temperate weather, and lower population density were associated with a decrease in R_t_. Of these county-specific factors, social distancing appeared to be the most significant in reducing SARS-CoV-2 transmission.

*Meaning:* R_t_ varies significantly across counties. The relationship between R_t_ and county-specific factors can inform policies to reduce SARS-CoV-2 transmission in selective and heterogeneous communities.

## Introduction

The coronavirus disease Covid-19 is the result of the novel severe acute respiratory syndrome coronavirus SARS-CoV-2. This virus has caused a pandemic resulting in over 3.2 million cases of Covid-19 by May 1st, 2020, with over 236,000 deaths worldwide. By the same date, there were over 1 million individuals with Covid-19 in the United States (US) resulting in over 64,000 deaths. The rapid evolution of this pandemic led to widespread implementation of social distancing measures across most of the US and the world.

The transmissibility of SARS-CoV-2, like other viral pathogens, is estimated by the reproduction number (R). An infectious pathogen’s R value represents the number of people that will be infected by an individual who has the infection. An R value that exceeds one will result in increasing numbers of incident cases as each infected individual will transmit the infection to more than one individual. When the R is below one the transmission of that pathogen will eventually cease as each infected patient will transmit infection to less than one person. Therefore, the R value is an important measure to estimate when attempting to predict the evolution of an outbreak. Often it is assumed that R is constant for each pathogen, however, the R most certainly varies by location and by time, referred to as the instantaneous R or R_t_. ^1-3^ Variation in R_t_ is likely dependent on community specific factors such as population density, temperature and/or humidity, employed policy change such as social distancing, and number of susceptible individuals.

Models that rely on fixed assumptions for R_t_ are therefore unlikely to capture local heterogeneity in transmission. Our objective was to understand how county-specific factors – population density, social distancing, temperature, and humidity – influence the R_t_, across counties in the US. Understanding how county-level factors impact SARS-CoV-2 R_t_ will allow policy makers to implement targeted interventions to decrease R_t_ in heterogeneous communities.

## Methods

### Setting and Participants

We selected 260 counties representing 55% of the total US population based on the following characteristics: 1) county with at least one case as of February 25, 2020; 2) county containing at least one city with population exceeding 100,000 persons; 2) county includes a state capital; or 3) most populated county in a state. We excluded counties with average daily case rates less than five and/or counties with fewer than 3 days of daily case rates exceeding five over the analysis period between February 25 and April 23, 2020. We considered time zero for each county the date at which they achieved the minimum threshold of disease activity. In total, 211 counties, representing 46 states and the District of Columbia, met these criteria.

### Outcome

The outcome is the estimated R_t_ of SARS-CoV-2 in each county. Daily incident case counts of Covid-19 aggregated at the county level were obtained from the *New York* Times.^4^

### Exposures

There were three *a priori* exposure variables of interest: social distancing practice, population density, and daily mean wet-bulb temperatures. Social distancing was measured using a dataset of daily cell-phone movement provided by Unacast.^5^ Based on an *a priori* assumption and confirmed by preliminary analyses that included cellphone measurement of overall distance traveled, we used a social distancing variable that measured percent change in visits to non-essential businesses (*e.g*., restaurants, hair salons) within each county, compared to visits in a four-week baseline period between February 10th and March 8th, 2020. We used a rolling average of the percentage of visits 3 to 14 days before time zero, based on the lag observed between change in social distancing and averaged R_t_ estimates across the counties (eFigure 1), and on an incubation period of at least 3 days.^6^

Population density of each county was obtained from US Census data and is expressed as number of people per square mile. Log transformation was performed to achieve a normal distribution because of significant skewness in density for the largest cities.

Wet-bulb temperatures are a metric that capture the thermodynamic relationship of temperature and humidity and have been applied in the study of weather and human mortality and other human disease.^7,8^ These measurements were obtained from National Oceanic and Atmospheric Administration (NOAA) Local Climatological Data.

### Covariates

County-level covariates that may confound the association between exposures of interest and R_t_ were considered, and included demographic factors (*e.g*. age distribution, insurance status and socioeconomic status), and health-related factors associated with Covid-19 severity (e.g. proportion of individuals with hypertension, obesity or diabetes, and proportion of smokers).^9^ Demographic and health characteristics were abstracted from the US Census, American Community Survey, Behavioral Risk Factor Surveillance System, Esri Business Analyst, and Multi-Resolution Land Characteristics Consortium.

We identified ten clinically meaningful covariates, examined the correlation between each pair of factors, and calculated the variance inflation factor to quantify multicollinearity among variables (eFigure 2). Among highly correlated variables, we chose covariates based on their potential relationship with viral transmission and/or the probability of an infected individual becoming symptomatic and therefore obtaining a diagnostic test. The final covariates included proportion of residents over age 65 years, those <200% of poverty level, and percentage of the population with diabetes. Variables for obesity, smoking, and uninsured population were removed due to collinearity with other variables.

### Statistical Analysis

Our analysis followed a two-step procedure. First, we calculated the R_t_ for SARS-CoV-2 using the method of Cori *et al*. with a moving average window of 3 days.^1^ This method has been applied in the dynamic estimation of R_t_ from Wuhan.^10^ The generation time of SARS-CoV-2 was assumed to follow a gamma distribution with mean of 7.5 days and standard deviation of 3.4 days according to a previous epidemiological survey of the first 425 cases in Wuhan.^6^ In the early days of a county’s outbreak, when the ratio of cases/tests was unstable, kernel smoothing with a box kernel and bandwidth of 7 days was performed to account for the likelihood that cases from prior days would accumulate when testing capacity increased.^11,12^ We stopped smoothing one day after the county’s daily cases/tests ratio first fell in the interval of [5%, 90%] or after March 20, 2020 to avoid over-processing the data.

Next, we fit a linear mixed effects model with a random intercept for each county to evaluate the association between exposures and R_t_ after a log transformation, adjusting for covariates. Population density was standardized after log transformation due to the highly skewed distribution. Temperature effects were estimated using a distributed lag non-linear model (DLNM), which consider bi-dimensional exposure-lag-response relationships between wet bulb temperature and log(R_t_).^13-15^ We considered a lag period of 4- to 10-days before case identification to reflect the incubation period of SARS-CoV-2 and to reduce bias introduced by daily weather influencing the decision for an individual to seek a test. The final cross-basis term included natural cubic splines defined by 3 internal knots at the 10^th^, 75^th^, and 90^th^ percentile of temperature ranges observed during the period, corresponding to 33°F and 55°F, and 67°F, and 2 knots in the lag dimension at 6 and 8 lag days. The number and placement of spline knots were based on an *a priori* assumption of a relatively simple relationship between temperature and SARS-CoV-2 transmission and on minimization of the Akaike information criterion. The temperature knots permitted flexibility in the model nonlinearity at higher and lower temperatures.^16^ We included an interaction between population density temperature, assuming that temperatures would influence transmissibility differently in densely populated counties.^17^ The relative change in R_t_ was expressed as the cumulative-exposure-response relative to 53°F. Interactions between population density and social distancing were included in the linear mixed effects model, given the hypothesis that the impact of social distancing might be greater in highly dense areas.

### Sensitivity Analyses

First, the model was re-estimated every two weeks (or 3 times) over a period of one month, checking for stability in estimates of effects for primary covariates. Second, the model fit was evaluated by calculating in-sample R-squared (R^2^) in randomly selected 70 percent of the counties over 100 replicates. Finally, we assessed alternative temperature metrics, including dry-bulb temperatures, relative humidity, and absolute humidity; given the similarity of findings, we report the wet-bulb temperatures due to their simplicity in interpretation (data not shown).

Using publicly de-identified data, this study was exempt from IRB Review. Analyses were performed with R (version 3.6.0; R Development Core Team) using the *EpiEstim* and *dlnm* packages.^18,19^

## Results

Geographic locations and characteristics of the 211 counties are demonstrated in Figure 1 and Table 1. The 21 counties in the top decile for population density had the highest incident case and fatality rate per 100,000 people, nearly 10 times the estimates in the lowest quartile. Average R in the first two weeks was 5.7 (SD 2.5) in the top decile, compared to 3.1 (SD 1.2) in the lowest quartile. The average change in visits to non-essential businesses was higher among counties in the top decile compared to those in the lowest quartile (78% vs. 66%). The top decile of counties also experienced colder temperatures during the analysis period.

**Figure 1.**
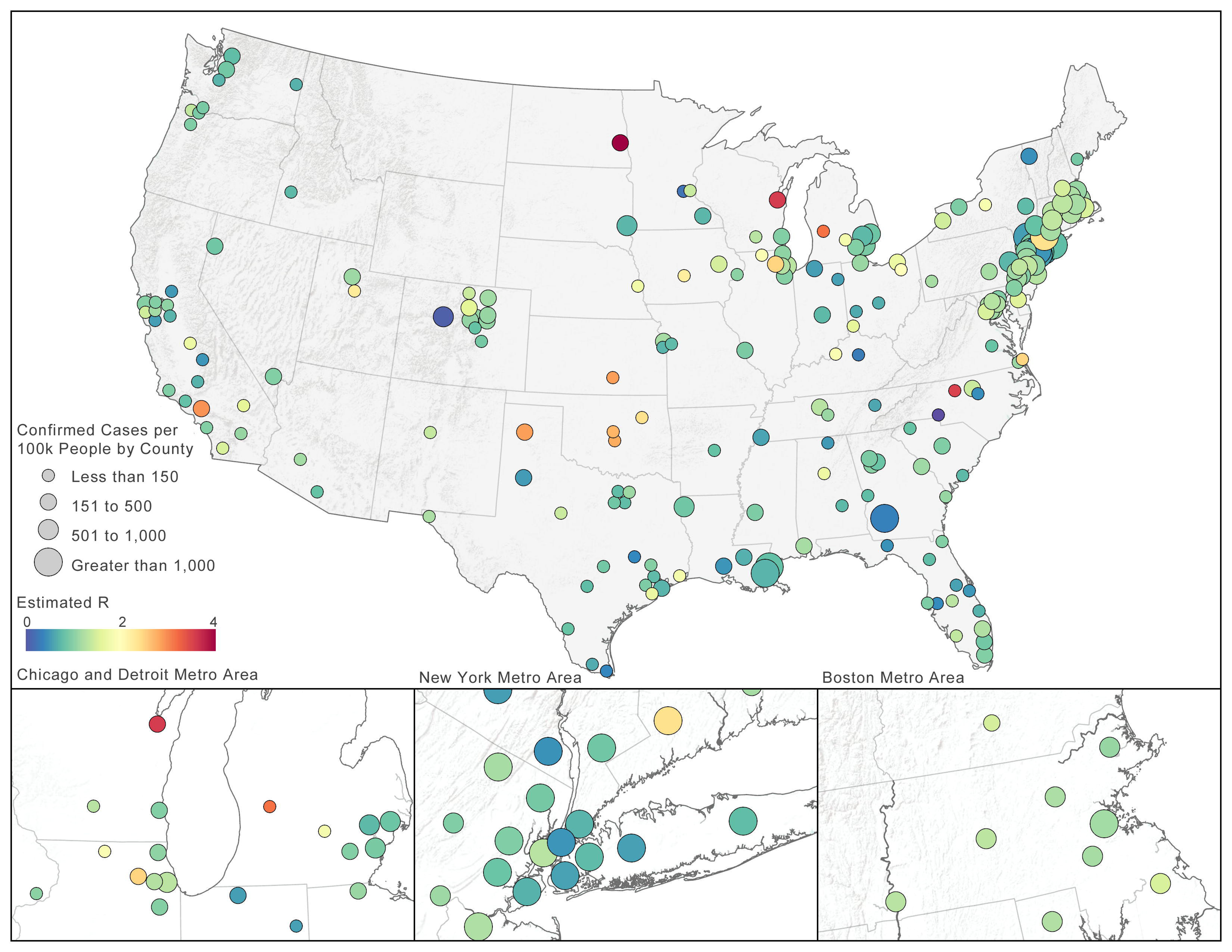
Location and estimated R_t_ of SARS-CoV-2 as of April 26, 2020 in 211 Counties in the United States. SARS-CoV-2 incidence per 100,000 persons as of April 26, 2020 are demonstrated by the relative size of the county locators. As of April 26, 2020, 211 counties in the United States were included in the analysis.

**Table 1:** Population Characteristics in an Analysis of Instantaneous R_t_ for SARS-CoV-2, by Population Density, for 211 Counties Across the United States.

|  | Population Density |  |  |  |  |  |
| --- | --- | --- | --- | --- | --- | --- |
|  | 0 to 25th percentile<br>(N = 53 counties) | 25th to 50th percentile<br>(N = 53 counties) | 50th to 75th percentile<br>(N = 53 counties) | 75th to 90th percentile<br>(N = 31 counties) | >90th percentile<br>(N = 21 counties) | All (N = 211 counties) |
| **Characteristics<sup>a</sup>** |  |  |  |  |  |  |
| Population density, median (IQR), per mi<sup>2</sup> | 298.4 (164.2 - 376.1) | 662.7 (539.1 - 826.8) | 1,368.7 (1,211.4 - 1,584.9) | 2,395.4 (2,151.2 - 2,860.6) | 8,916 (5,381.8 - 14,475.6) | 1,022.7 (471.2 - 1,846.0) |
| Hypertension, mean (SD), % | 29.4 (5.3) | 31.6 (3.9) | 31.2 (3.1) | 31.5 (3.8) | 29.3 (3.5) | 30.7 (4.2) |
| Diabetes, mean (SD), % | 9.2 (2.3) | 10.1 (1.8) | 10.0 (1.3) | 10.3 (1.5) | 10.1 (1.5) | 9.9 (1.8) |
| Regular smokers, mean (SD), %<sup>b</sup> | 16.5 (2.4) | 16.9 (2.3) | 16.3 (2.6) | 16.6 (2.7) | 15.9 (2.5) | 16.5 (2.5) |
| BMI > 30, mean (SD), % | 29.6 (4.5) | 30.9 (3.5) | 30.2 (3.5) | 31.1 (3.8) | 28.2 (4.1) | 30.1 (3.9) |
| Age <18 years, median (IQR), % | 23.9 (22.1 - 25.7) | 23.4 (21.6 - 24.8) | 22.3 (21.4 - 23.8) | 23.1 (22.1 - 24.1) | 20.9 (18.3 - 22.2) | 22.7 (21.4 - 24.5) |
| Age 18 to 34 years, median (IQR), % | 24.5 (23.1 - 26.4) | 23.7 (22.0 - 26.1) | 23.3 (21.3 - 25.0) | 24.4 (22.7 - 25.9) | 28.3 (23.8 - 30.6) | 24.1 (22.3 - 26.2) |
| Age 35 to 64 years, median (IQR), % | 37.1 (35.5 - 38.6) | 38.1 (36.5 - 39.8) | 39.5 (38.1 - 41.0) | 38.9 (37.6 - 40.3) | 38.8 (37.1 - 40.6) | 38.4 (36.9 - 40.1) |
| Age ≥ 65 years, median (IQR), % | 13.8 (11.9 - 14.8) | 13.5 (12.6 - 15.2) | 14.7 (12.8 - 16.2) | 13.5 (11.8 - 14.7) | 13.2 (11.9 - 14.8) | 13.8 (12.4 - 15.4) |
| Low Income, mean | 33.2 (9.8) | 31.5 (8.6) | 27.0 (8.8) | 30.4 (8.5) | 31.2 (10.2) | 30.6 (9.3) |
| (SD), %<sup>c</sup> |  |  |  |  |  |  |
| Uninsured, mean (SD), % | 9.6 (4.7) | 9.5 (5.4) | 8.2 (4.1) | 9.4 (4.4) | 8.5 (3.1) | 9.1 (4.6) |
| Change in visits to non-essential businesses 2/24/20-3/8/20, mean (SD), %<sup>d</sup> | -2.4 (7.0) | -2.5 (6.1) | -3.0 (5.7) | -3.1 (6.1) | -2.8 (6.3) | -2.7 (6.3) |
| Change in visits to non-essential businesses 4/6/20-4/19/20, mean (SD), %<sup>d</sup> | -66.3 (7.8) | -65.3 (7.3) | -69.2 (6.8) | -71.3 (5.8) | -77.9 (5.2) | -68.7 (7.9) |
| Daily wet bulb temperature, median (IQR), °F<sup>e</sup> | 46.3 (37.6 - 54.8) | 46.7 (39.4 - 58.6) | 46.8 (39.6 - 58.1) | 46.0 (39.0 - 56.6) | 42.6 (38.1 - 47.2) | 45.5 (38.8 - 55.0) |
| **Outcomes** |  |  |  |  |  |  |
| R in the first two weeks, mean (SD) | 3.1 (1.2) | 3.1 (1.1) | 3.6 (1.1) | 4.0 (1.6) | 5.7 (2.5) | 3.6 (1.6) |
| Cases per 100,000 people on 4/26/20, median (IQR) | 121.4 (87.8 - 175.4) | 126.7 (79.3 - 180.3) | 196.0 (73.1 - 530.4) | 206.6 (110.5 - 483.2) | 1,185.2 (313.2 - 1,891.2) | 154.7 (87.9 - 350.6) |
| Deaths per 100,000 people on 4/26/20, median (IQR) | 4.2 (1.9 - 8.0) | 4.2 (2.3 - 7.5) | 5.6 (2.3 - 28.6) | 8.9 (4.1 - 21.9) | 43.7 (10.4 - 106.7) | 5.8 (2.5 - 16.3) |

Abbreviations: SD, standard deviation; IQR, interquartile range; BMI, body mass index (calculated as weight in kilograms divided by height in meters squared);
<sup>a</sup> All characteristics were obtained from the American Community Survey (2018) except health data, which were obtained from the CDC Behavioral Risk Factor Surveillance System (2017), social distancing obtained from Unacast (2020), and wet bulb temperature from National Oceanic and Atmospheric Administration (2020).
<sup>b</sup> Regular smokers was defined as adult respondents who reported smoking $\geq 100$ cigarettes in their life and currently smoke at least some days.
<sup>c</sup> Low income was defined as $<200\%$ poverty level
<sup>d</sup> Visits to non-essential businesses obtained from Unacast, calculated as the change from the average non-essential business visits during matching days of the week before 3/9/2020.
<sup>e</sup> Daily wet-bulb temperatures were calculated by averaging the hourly recordings from weather stations that contribute to the National Oceanic and Atmospheric Administration's Local Climatological Data

R_t_ was estimated over a total of 6588 county days for the 211 counties; 17 outlier county days were removed due to an extremely low estimate of R_t_ at low case thresholds (R_t_<0.05). The estimated R_t_ by county varied greatly but on average had a high elevation earlier the epidemic, reaching a peak R_t_ of 7.8 before declining later in the period (eFigure 1). Adjusting for county-level covariates, social distancing, population density, and temperature were significantly associated with R_t_ (p<0.001; Table 2). A 50% decrease in visits to non-essential businesses was associated with a 57% decrease in R_t_ (95% CI, 56% to 58%). Compared to counties with population density at the 25^th^ percentile, the 20 counties in the top decile of density had a 16% increase in the relative R_t_ (95% CI, 9% to 27%).

**Table 2.**
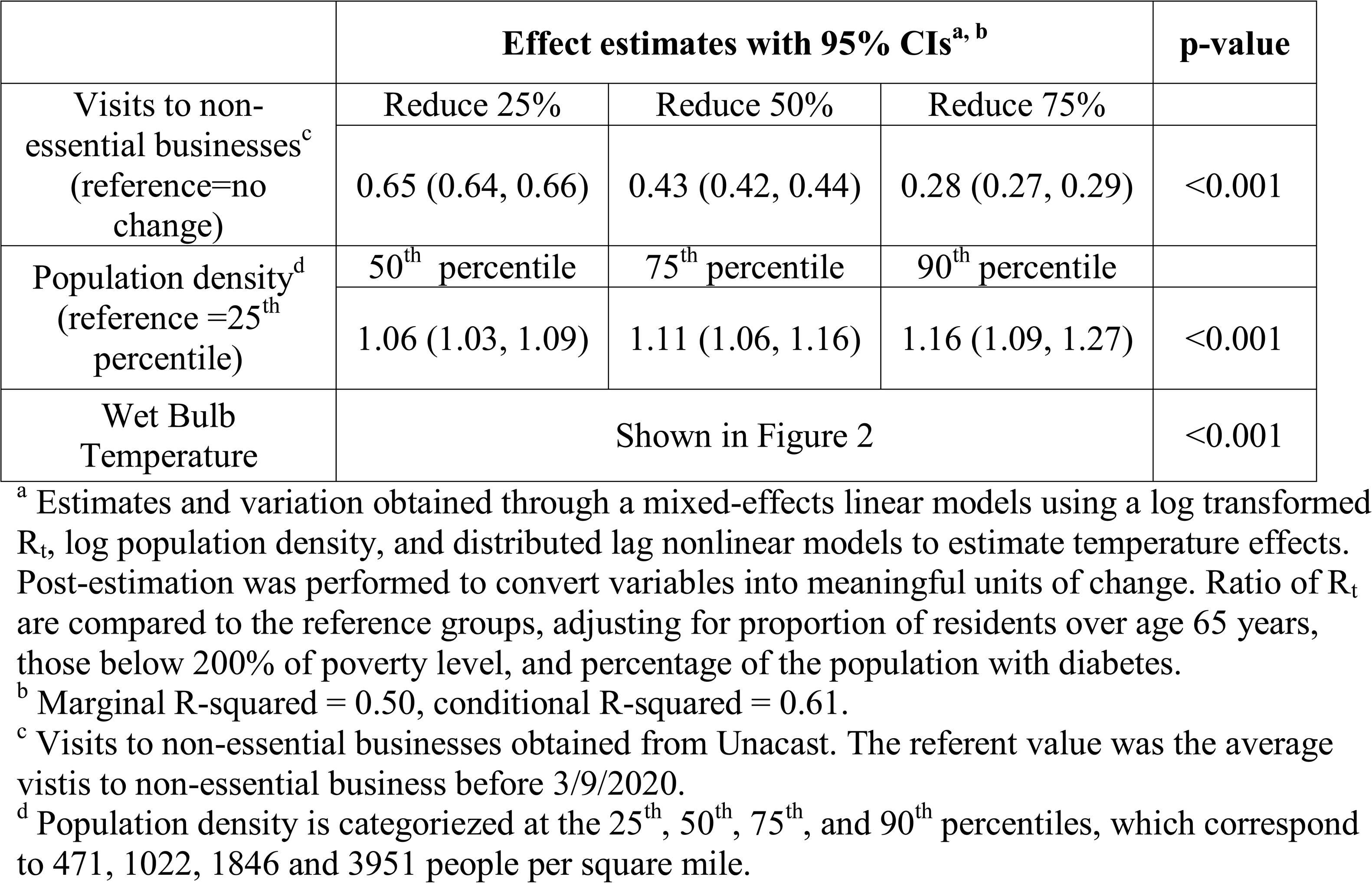
Ratios of R_t_ for Social Distancing and Population Density in 211 United States Counties between February 25 and April 23, 2020.

|  | Effect estimates with 95% CIs<sup>a, b</sup> |  |  | p-value |
| --- | --- | --- | --- | --- |
| Visits to non-essential businesses<sup>c</sup><br>(reference=no change) | Reduce 25% | Reduce 50% | Reduce 75% |  |
|  | 0.65 (0.64, 0.66) | 0.43 (0.42, 0.44) | 0.28 (0.27, 0.29) | <0.001 |
| Population density<sup>d</sup><br>(reference =25<sup>th</sup> percentile) | 50<sup>th</sup> percentile | 75<sup>th</sup> percentile | 90<sup>th</sup> percentile |  |
|  | 1.06 (1.03, 1.09) | 1.11 (1.06, 1.16) | 1.16 (1.09, 1.27) | <0.001 |
| Wet Bulb Temperature | Shown in Figure 2 |  |  | <0.001 |

<sup>a</sup> Estimates and variation obtained through a mixed-effects linear models using a log transformed $R_t$ , log population density, and distributed lag nonlinear models to estimate temperature effects. Post-estimation was performed to convert variables into meaningful units of change. Ratio of $R_t$ are compared to the reference groups, adjusting for proportion of residents over age 65 years, those below 200% of poverty level, and percentage of the population with diabetes.
<sup>b</sup> Marginal R-squared = 0.50, conditional R-squared = 0.61.
<sup>c</sup> Visits to non-essential businesses obtained from Unacast. The referent value was the average visits to non-essential business before 3/9/2020.
<sup>d</sup> Population density is categorized at the 25<sup>th</sup>, 50<sup>th</sup>, 75<sup>th</sup>, and 90<sup>th</sup> percentiles, which correspond to 471, 1022, 1846 and 3951 people per square mile.

The nonlinear association of lagged temperature effects between 4 and 10 days is reported in Figure 2. Compared to the minimum estimated R_t_ at 53°F, relative R_t_ increased across the coldest temperatures, with a maximum ratio of 1.5 (95% CI 1.30-1.73) at 32°F. A second, smaller peak of the relative ratio R_t_ to 1.28 (95% CI 1.12-1.46) was estimated at 66°F, before declining again at higher temperatures. Given concerns for limited representation of temperatures at the upper and lower ranges across counties of different population density, we did not include interaction effects between population density and temperature.

**Figure 2.**
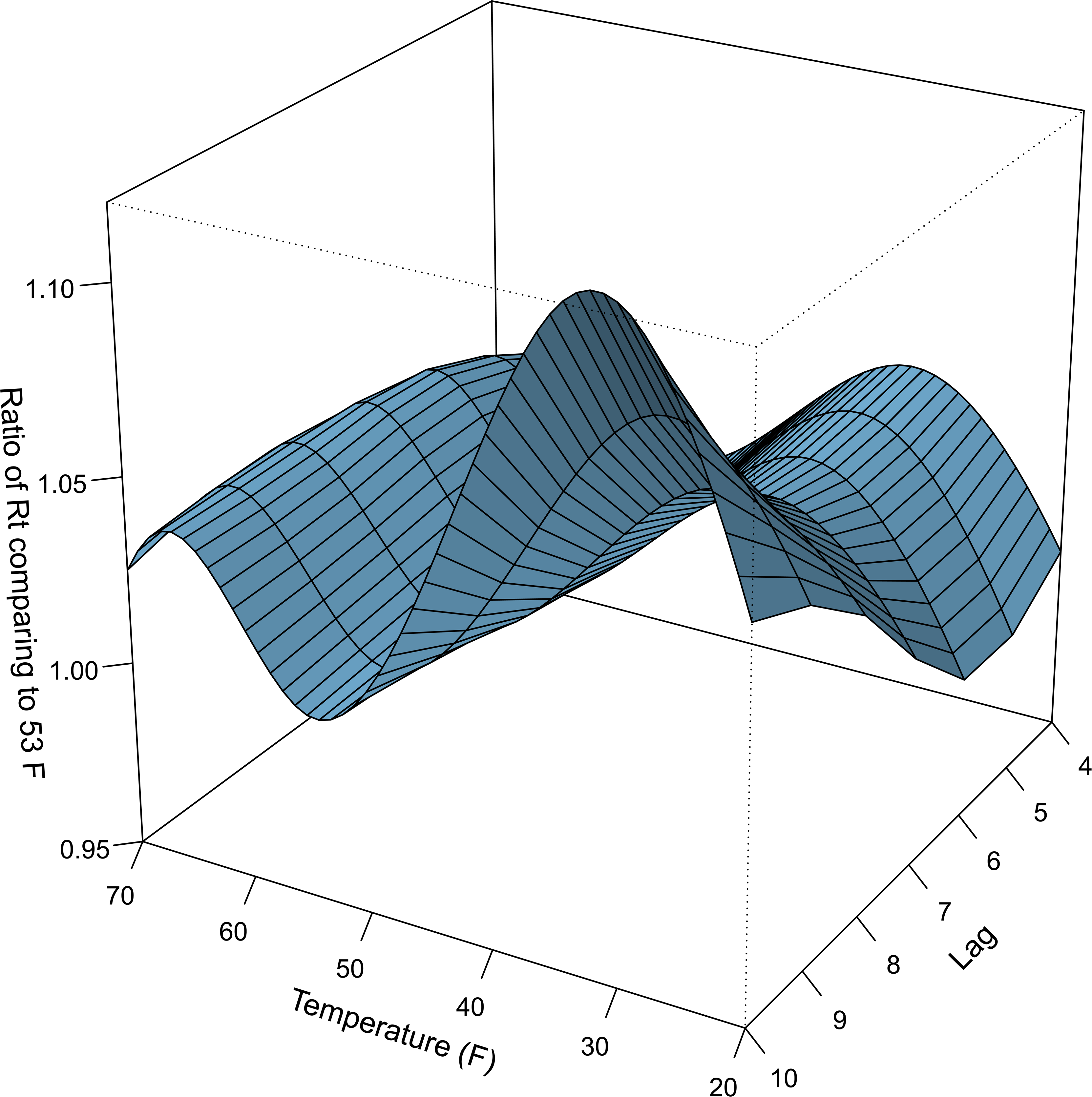
Cumulative Temperature-Dependence of SARS-CoV-2 R_t_ over a 7-day Lag Period (4 to 10 days) in 211 US Counties. A) The three-dimensional relationships of wet-bulb temperature (x-axis), lag (z-axis) and R_t_ for SARS-CoV-2 (y-axis) are shown. The point estimate of R_t_ at each point along the temperature range and lag window is shown, relative to 53°F. B) Cumulative exposure-response relationship between mean daily wet-bulb temperatures and R_t_ over a lag period of 4 to 10 days before case identification. The wet-bulb temperature range for the counties included in the analysis was 16°F to 77°F.

The standardized effect of social distancing, population density, and temperature on R_t_, appears in Figure 3. Assuming social distancing of 35% (or halfway between estimates during the US shelter in place phase and normal activity), 1% of counties were estimated to have a R_t_ less than 1.0 at 35°F, and 20% of counties were estimated to have a R_t_ of less than 1.0 at 55°F. When visits to non-essential businesses were reduced to the national average of 70%, the proportion of counties estimated to have R_t_ less than 1.0 increased to 49% and 96% at 35°F and 55°F, respectively. At this 70% reduction in visits to non-essential businesses, 62% and 98% of all counties in the lowest quartile of density achieved this threshold at 35°F and 55°F, respectively, compared to 5% and 86% of counties in the top decile at the same temperatures.

**Figure 3.**
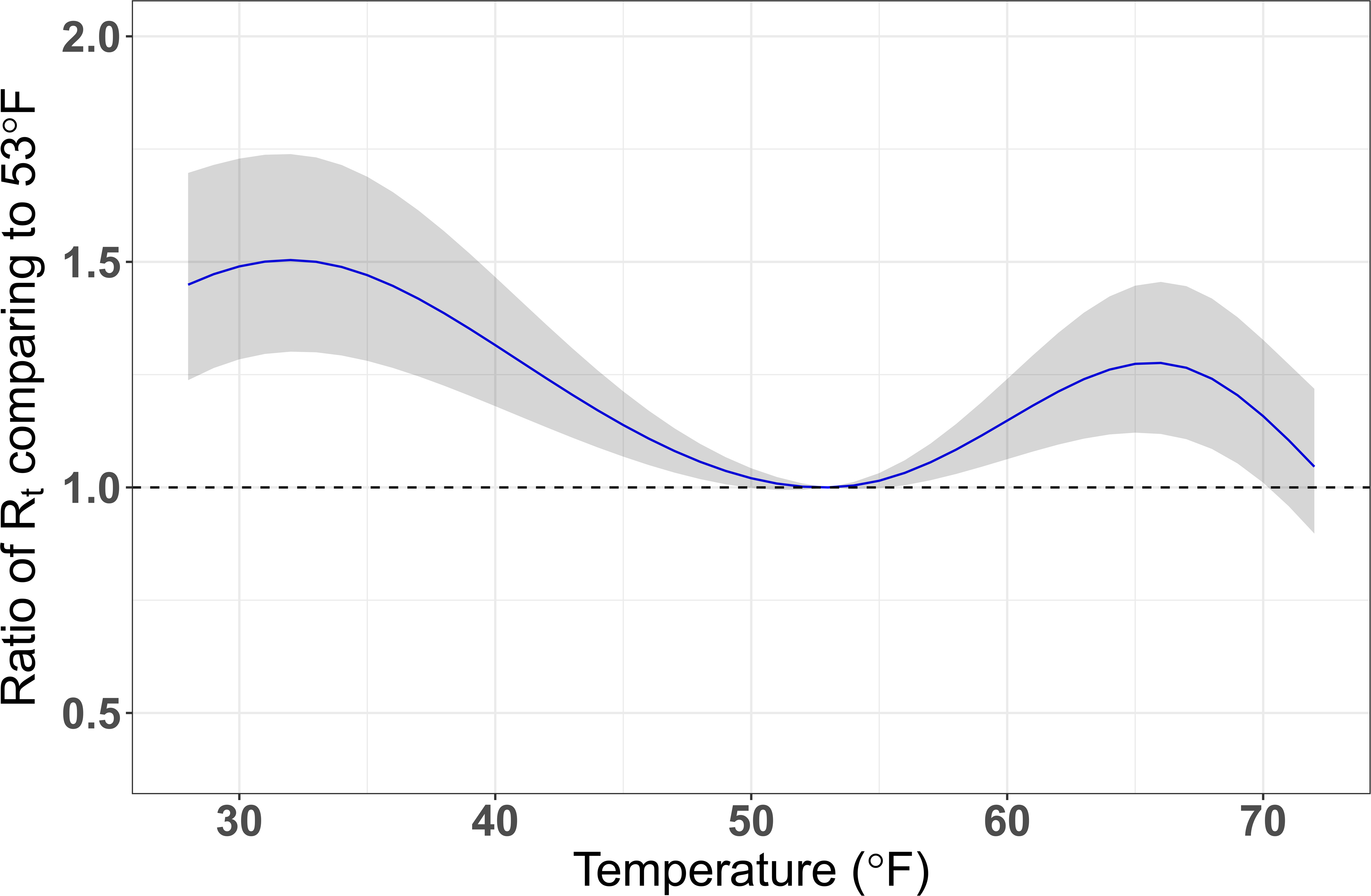
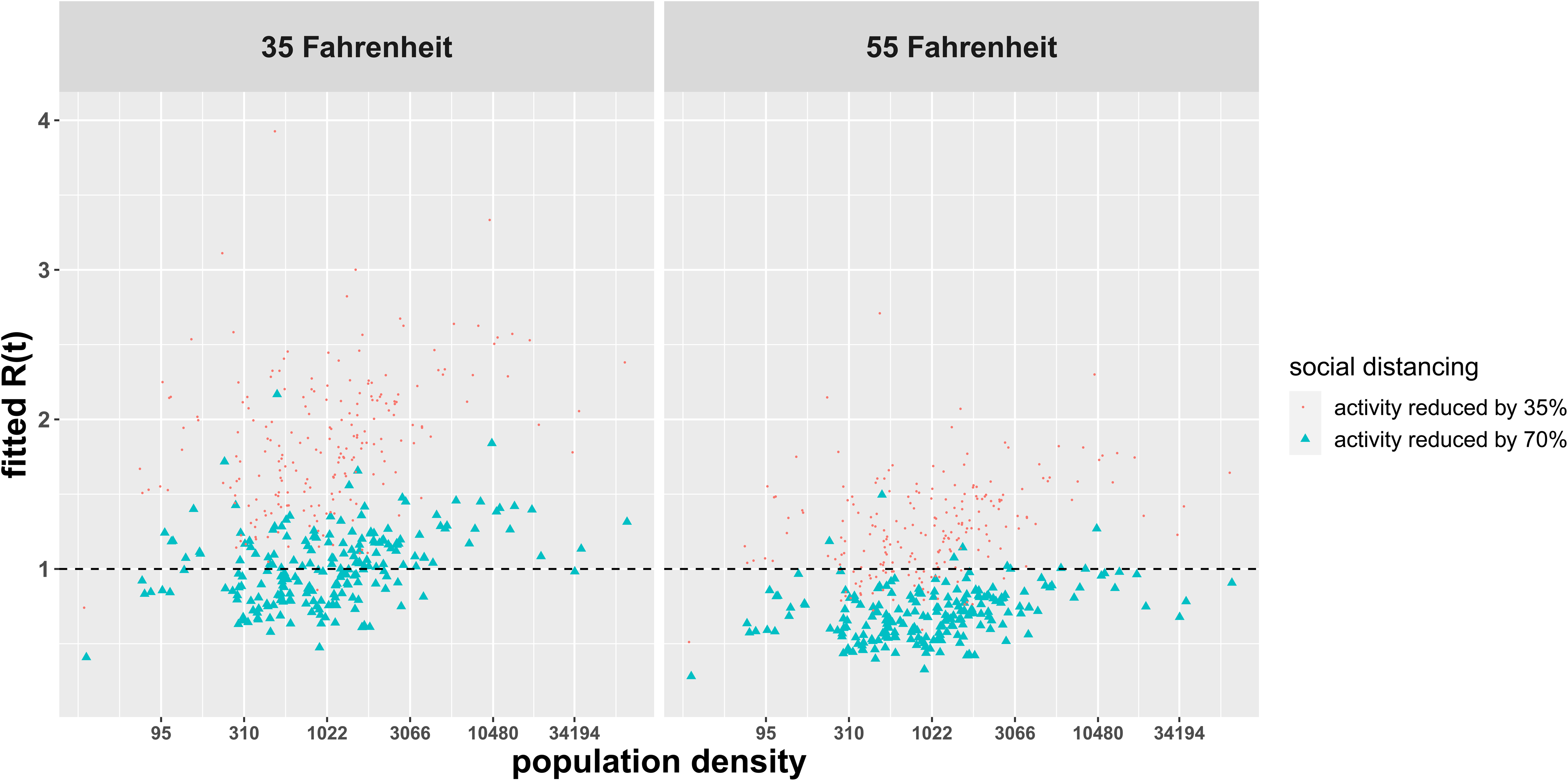
The Relative Impact of Social Distancing, Population density, and Temperature on SARS-CoV-2 R_t_ in 211 US Counties. Each point represents the fitted R_t_ of an individual county at 35°F (left panel) and 55°F (right panel), adjusted for the proportion of residents over age 65 years, those below 200% of poverty level, and percentage of the population with diabetes. Different levels of social distancing are shown for a 35% reduction (red circles) and 70% reduction (blue triangles) in visits to non-essential businesses.

No significant interactions were observed for population density and social distancing (p=0.7). Model estimates were stable over time. Overall R^2^ of the mixed effects model was 0.50 based on the covariate effects; the addition of random county intercepts increased the R^2^ to 0.61. Distribution of the in-sample R^2^ in randomly selected 70% of the counties over 100 replicates, revealing good model fit (eFigure 4).

## Discussion

In this analysis of 211 US counties, change in social distancing, population density, and temperatures were associated with the rate of SARS-CoV-2 transmission within a county, as measured by estimated R_t_. Our analysis indicates that of these three factors, implementation of social distancing has been the most significant in reducing transmission. While moderate temperatures were associated with lower R_t_, the mitigating impact of social distancing and temperatures were most important for more population dense counties, which had high R_t_ values that are consistent with higher R estimates from around the world.^20^

The underlying mechanism for the associations of social distancing and population density on the estimated R_t_ for SARS-CoV-2 are likely related to increased droplet transmission and potentially airborne transmission when individuals are in closer approximation to each other.^21,22^ However, the relationship of population density and Covid-19 outcomes may not be limited to transmission. The densest counties also had the highest number of deaths per 100,000 people. The association of more severe disease in higher density areas is hypothesized to be related to the inoculum effect. The inoculum effect suggests that individuals exposed to a higher viral load at the time of infection will have more severe illness, and is supported by epidemiologic studies for other viruses,^23,24^ and in particular for the SARS-1 coronavirus.^25,26,27^ These data support the concept that during a pandemic, people living in highly dense counties are more likely to transmit SARS-CoV-2 and to be exposed to higher inoculums of SARS-CoV-2. This translates not only to more cases of Covid-19, but also to a higher case fatality rate. Data assessing the inoculum effect for SARS-CoV-2 are needed to confirm this hypothesis.

Our analysis, which used well-established methods developed to examine effects of temperature on human health, is also among the first to consider the effects of temperature and humidity on SARS-COv-2 transmission.^13,28^ We found that R_t_ decreased with moderate temperatures, as temperatures increased from 32°F to 53°F. Beyond 53°F, there was a modest increase in R_t_ before R_t_ began declining again at higher temperatures. Our findings are consistent with the inverse relationship of temperature and transmission in animal models for influenza and other coronaviruses.^29,30^ Higher humidity was also found in those studies to increase viability and transmission of influenza through fomites, even as aerosol transmission is mitigated. If SARS-CoV-2 has similar properties to influenza at higher humidity it could explain our association of increased wet-bulb temperatures above 53°F with a modest increase in R_t_.

Beyond the direct effects of temperature and humidity on virus stability and viability, it is also possible that changes in these parameters alter human activity. For example, at higher temperatures people may more likely to congregate in public locations such as beaches and festivals. Therefore, an increase in R_t_ at higher temperatures could also be explained in part by a decrease in social distancing not measured by our social distancing variable. Regardless of the etiology for this association, we remain cautious in interpreting the effects of higher temperature and humidity on R_t_ beyond the temperate range we observe in this analysis. The coming months will allow for additional assessment of R_t_ during more prolonged periods of higher temperature and humidity. These additional observations will help to confirm or refute this association.

To date, projections of Covid-19 outcomes have considered large areas, such as countries, provinces, and US states. These models have provided useful information to set expectations for deaths in the US, identify potential gaps in health services such as ICU beds and ventilators, and guide initial wide-ranging social distancing recommendations from the federal and state governments. The value of our analysis at the county level has allowed us to better examine relevant contributions of social distancing, population density, and seasonal weather changes on a given county’s R_t_. This approach gives valuable information on risk of transmission to inform area-specific public policy decisions. It will be important to examine whether the introduction of these effects into models can predict—with some accuracy—over future weeks the likelihood of viral transmission as these predictive factors continue to change. To the degree that predictive modeling efforts are successful, it may demonstrate some validity of using this approach that incorporates random effects for counties alongside seasonal changes to inform future epidemics.

There are always limitations in observational studies. Generalizability remains a concern, particularly given our focus on larger counties. The 45% of the population not captured in our analysis were residing in smaller rural counties and as such our models are not applicable to these areas. Their omission likely attenuates the effects we observe, at least for population density. We considered total cases reported within each county. It is possible that differences in diagnostic test availability could contribute to the variation detected by the random effects across counties. However, our estimate of R_t_ depends on the rate of change of cases rather than the absolute number of cases, which should limit the impact diagnostic test availability. Further, we smoothed early outbreak case incidences to account for early limited access to diagnostic tests. We intentionally did not include testing capacity as a covariate, so as not to overfit the model (*e.g*., controlling for a factor that was also associated with rising viral transmission itself). As the random county intercept explained additional variation, there are likely unmeasured county factors that we did not capture. These factors might include commuter automobile traffic, public transportation usage, and domestic and international flights, which had decreased during the study period. It is clear that early local epidemics were seeded by international travel that contributed to early transmission in some locations.^31^ Further investigation will be needed as communities re-open to examine the impact of these additional time-varying factors on risk for SARS-CoV-2 transmission

## Conclusions

This explanatory analysis suggests that social distancing and differences in population density and temperatures account for variation in the R_t_ for SARS-COv-2. These results may guide local policy decisions for managing this pandemic in counties across the US.

## Data Availability

There is no availability to the data referred to in the manuscript.

## Acknowledgements

The authors have no relevant conflicts of interest, including financial interests, activities, relationships, and affiliations, that relate to the research described in this paper. The authors received no financial support for the research, authorship, and publication of this paper.

---

Online only supplements included in this document: eTable 1, eFigure 1, eFigure 2, eFigure 3

**eTable 1:**
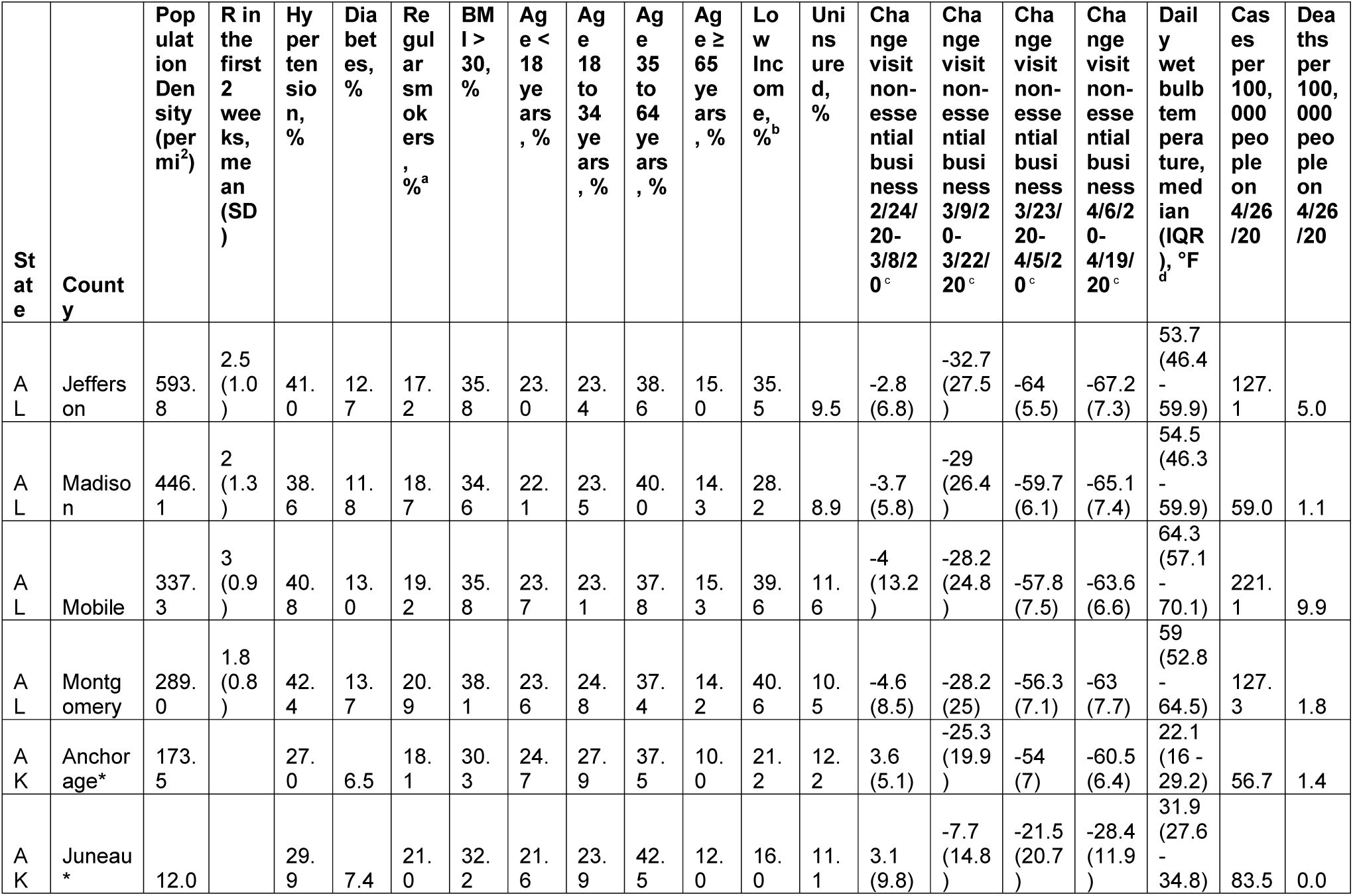
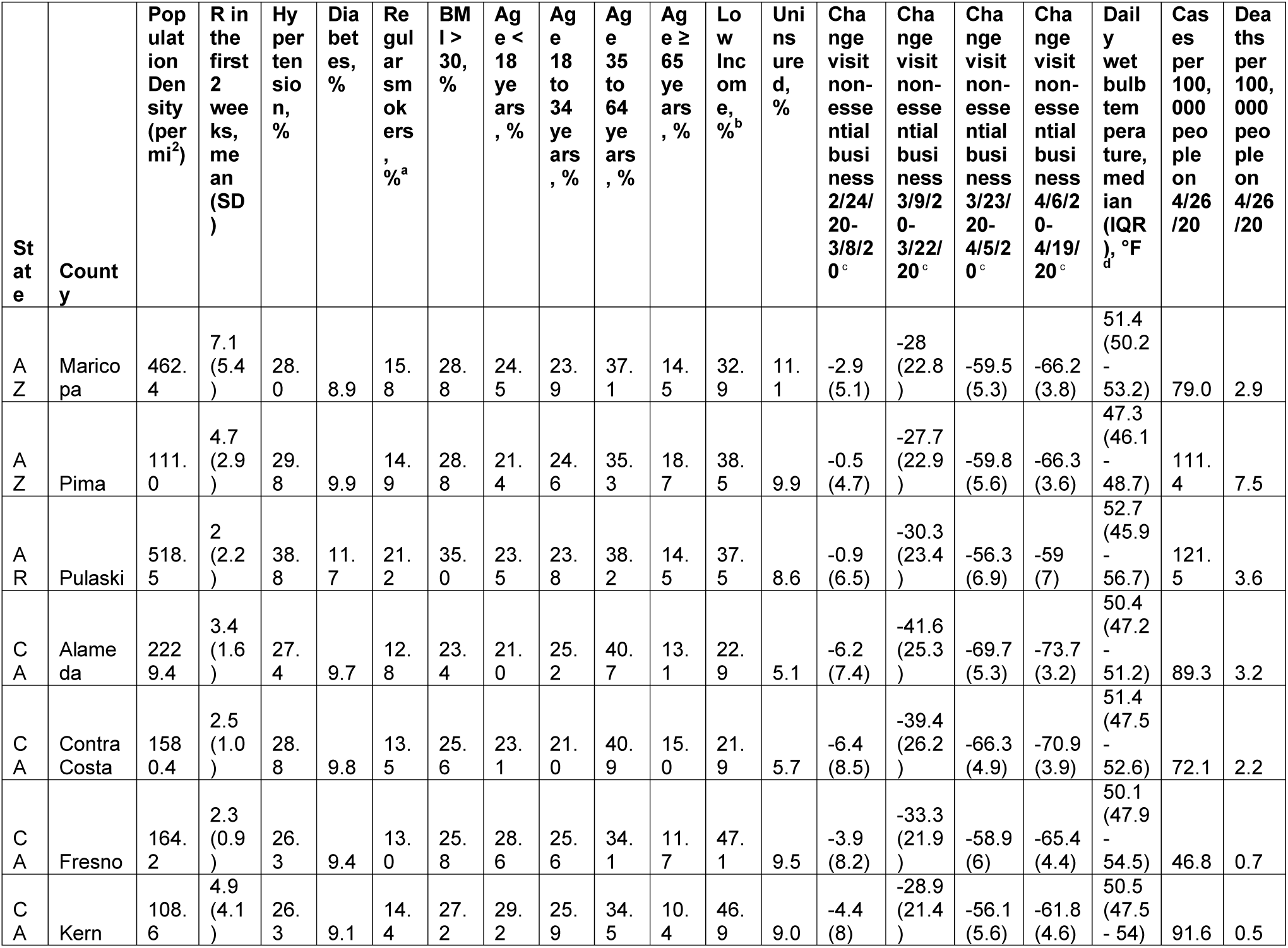
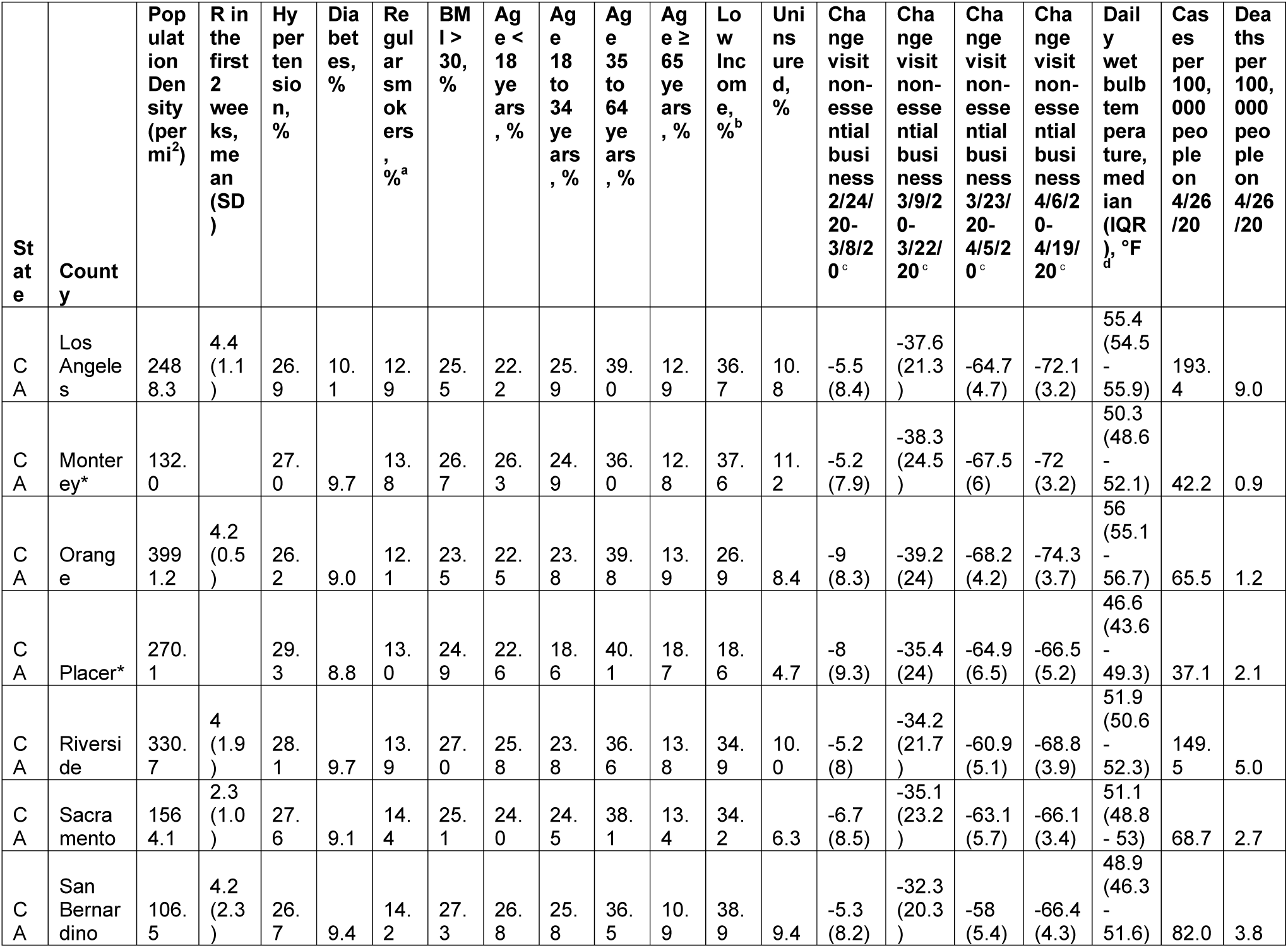
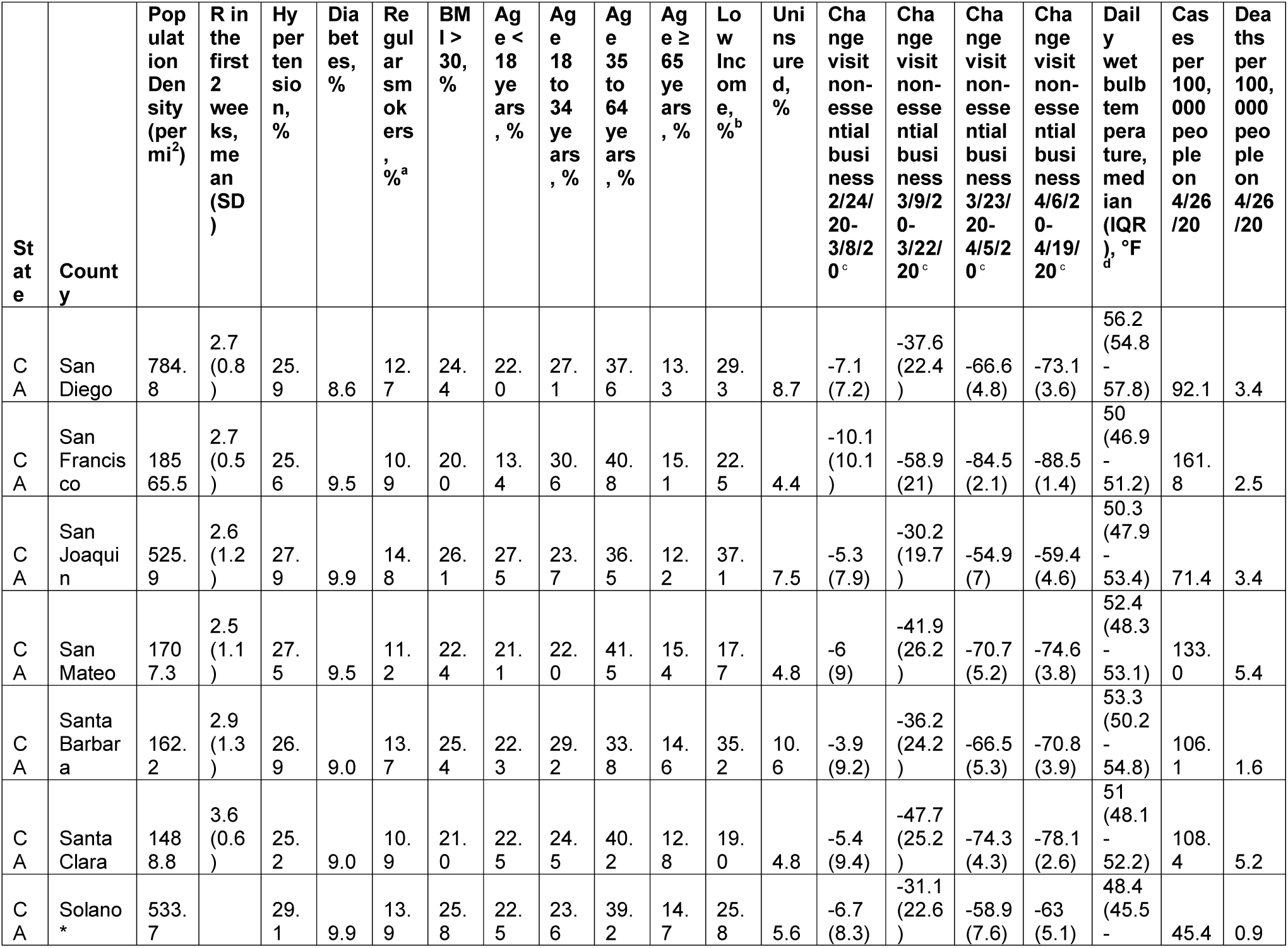
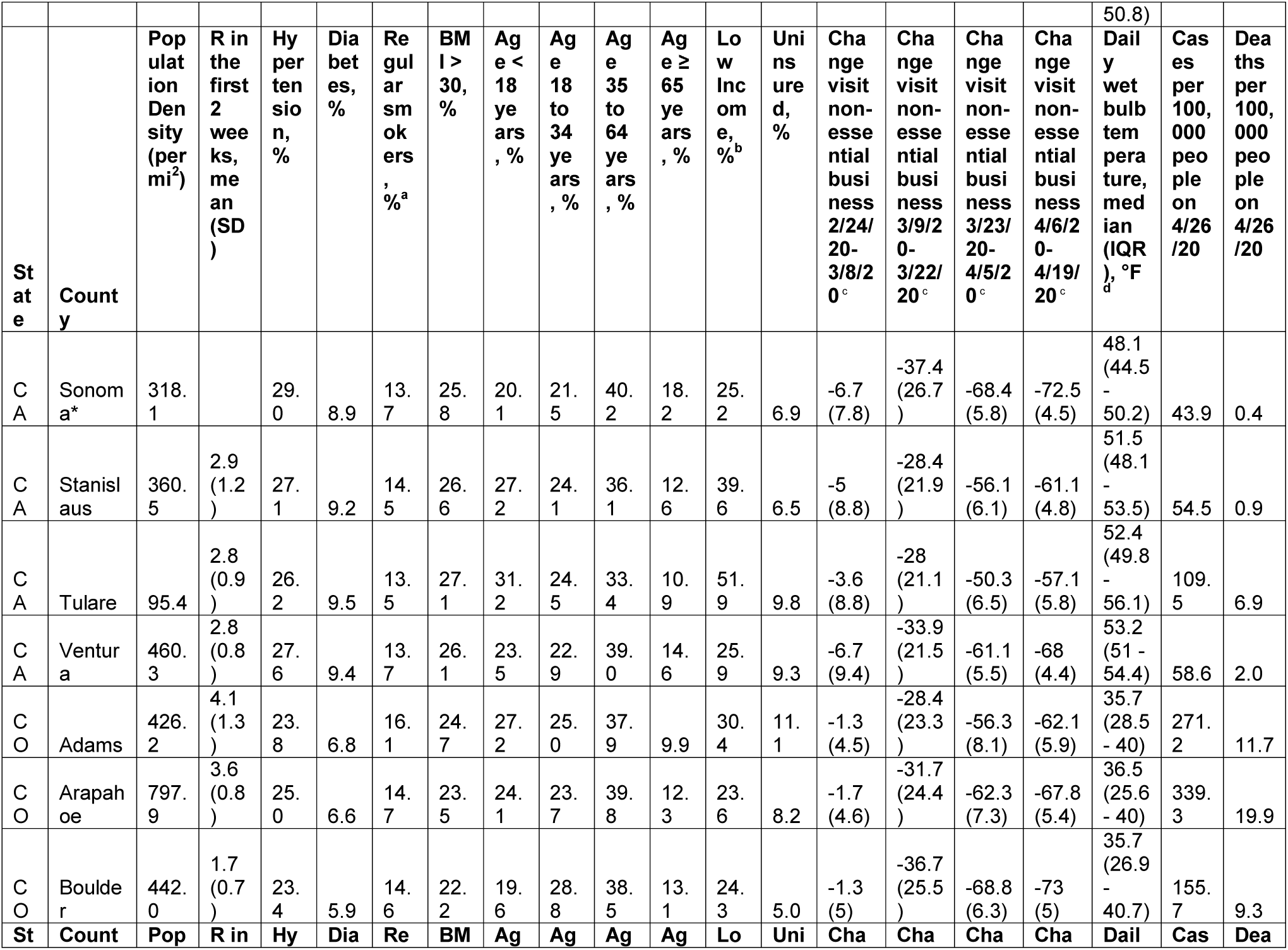
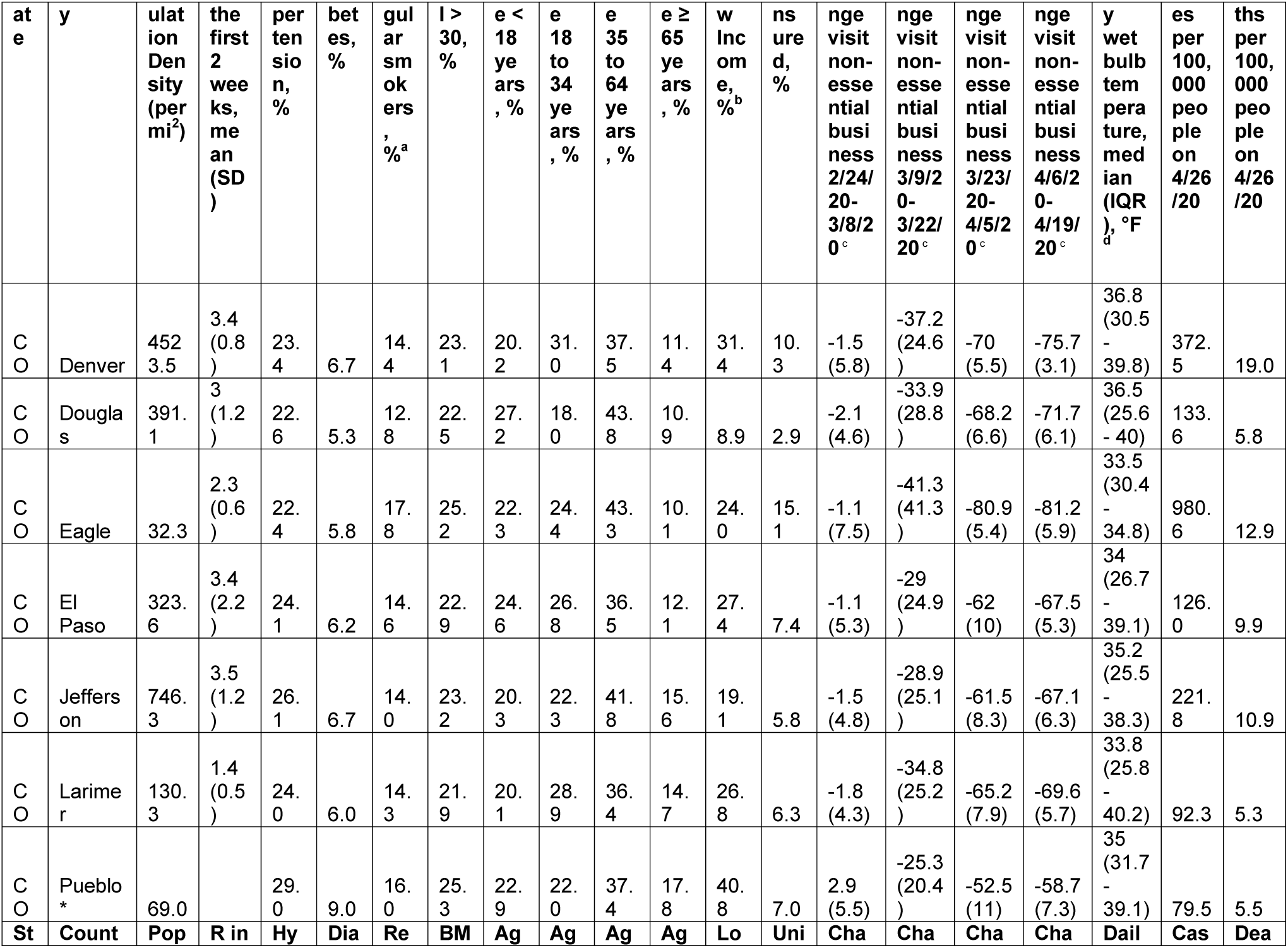
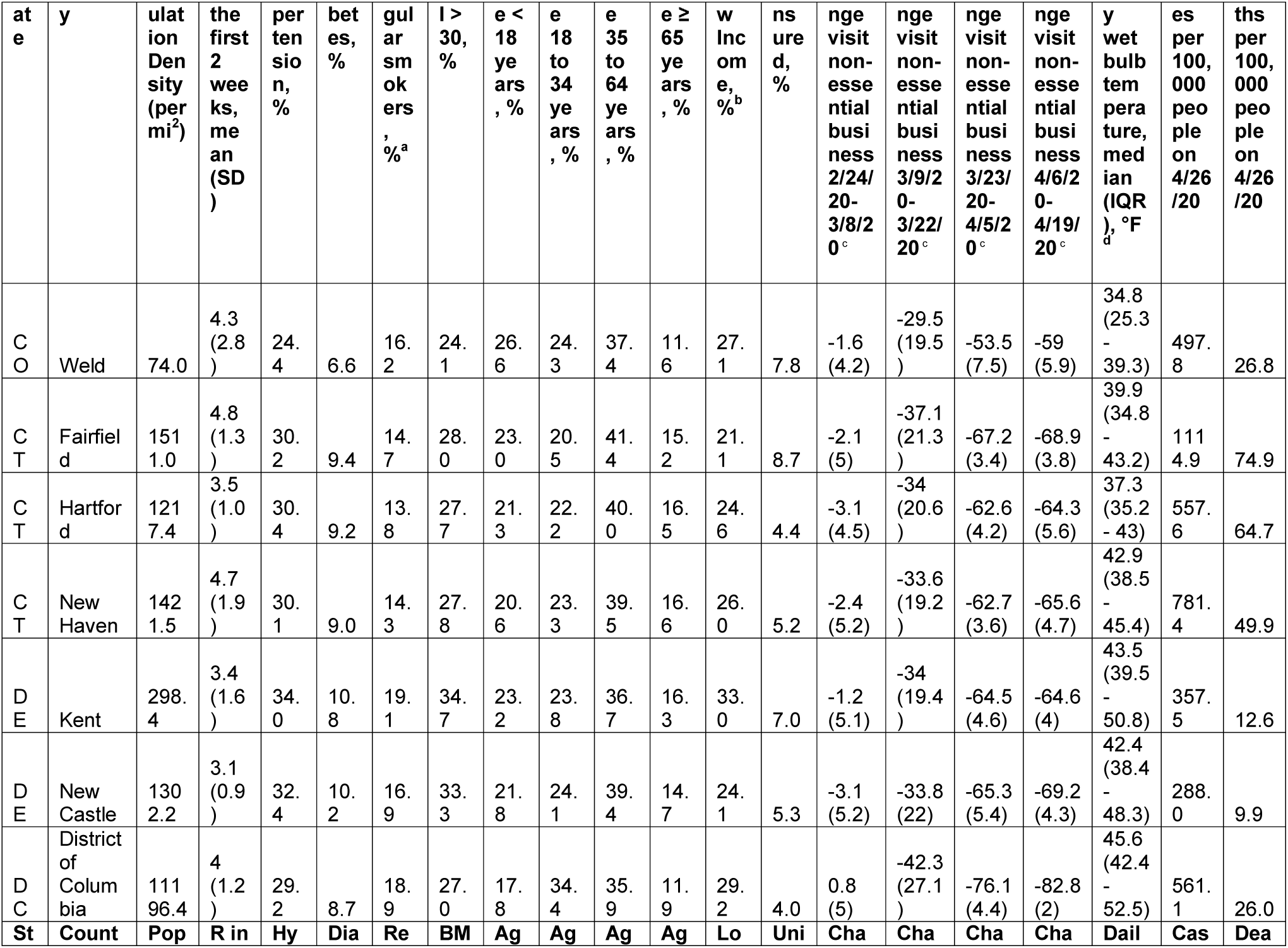
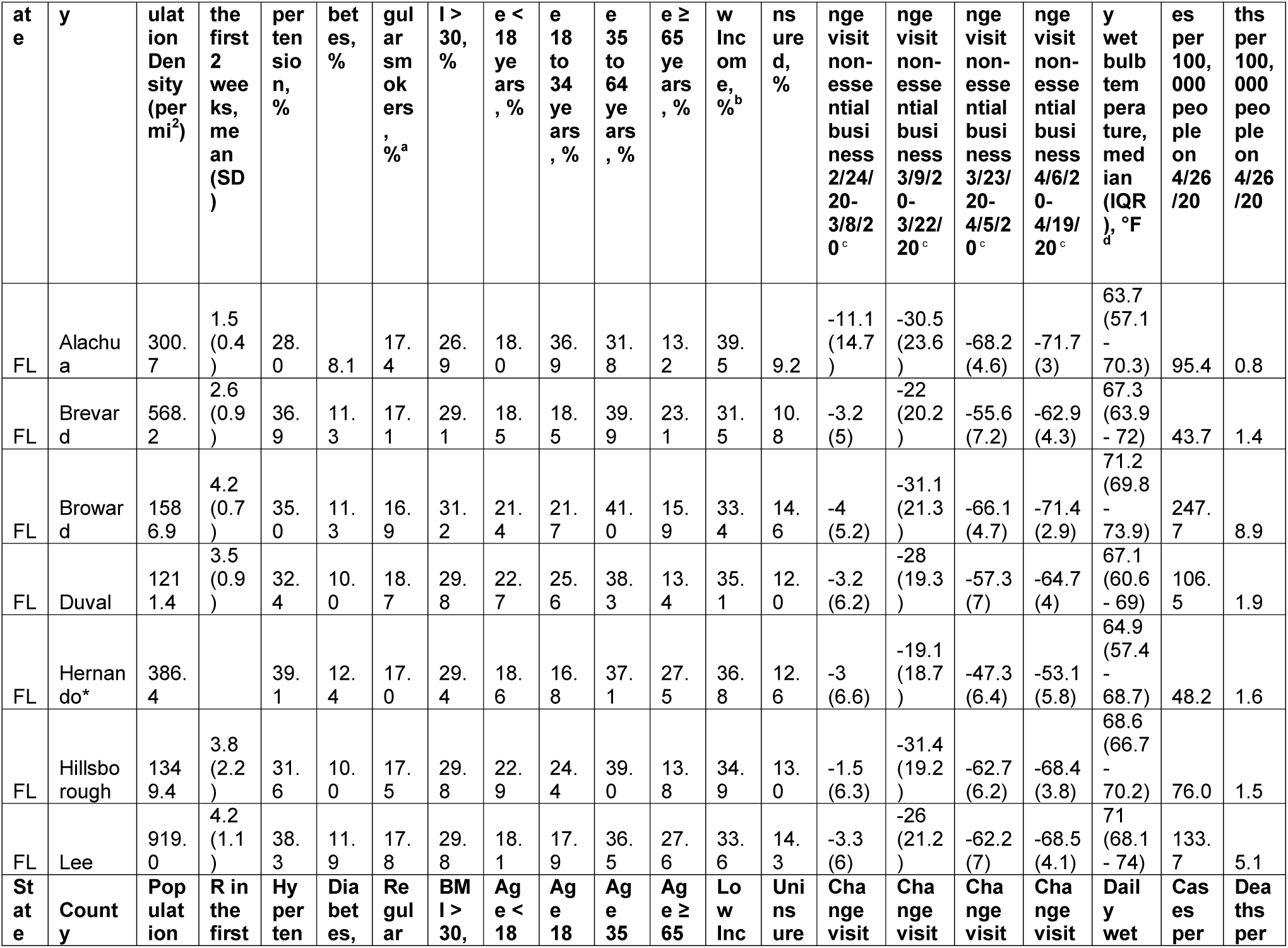
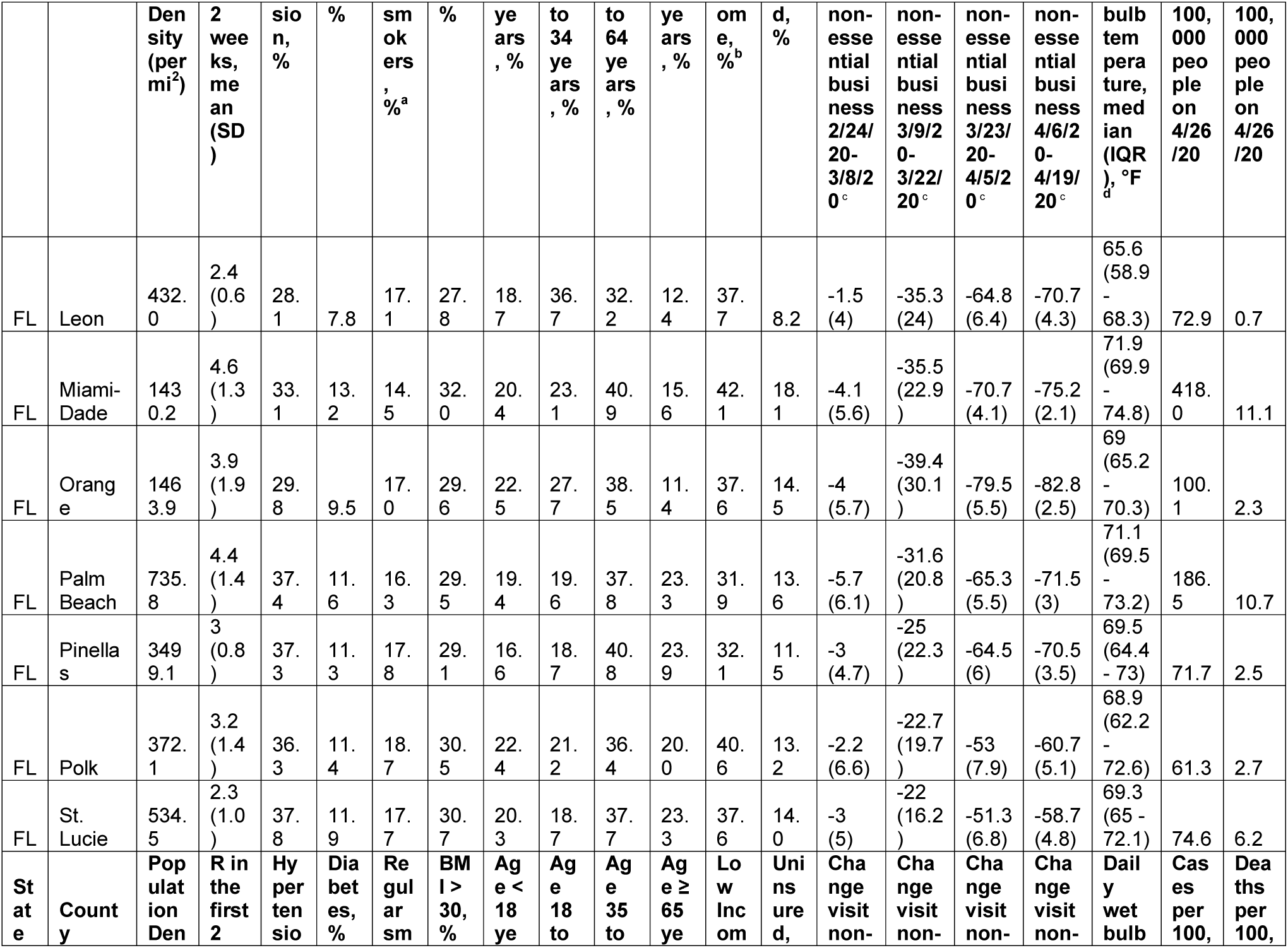
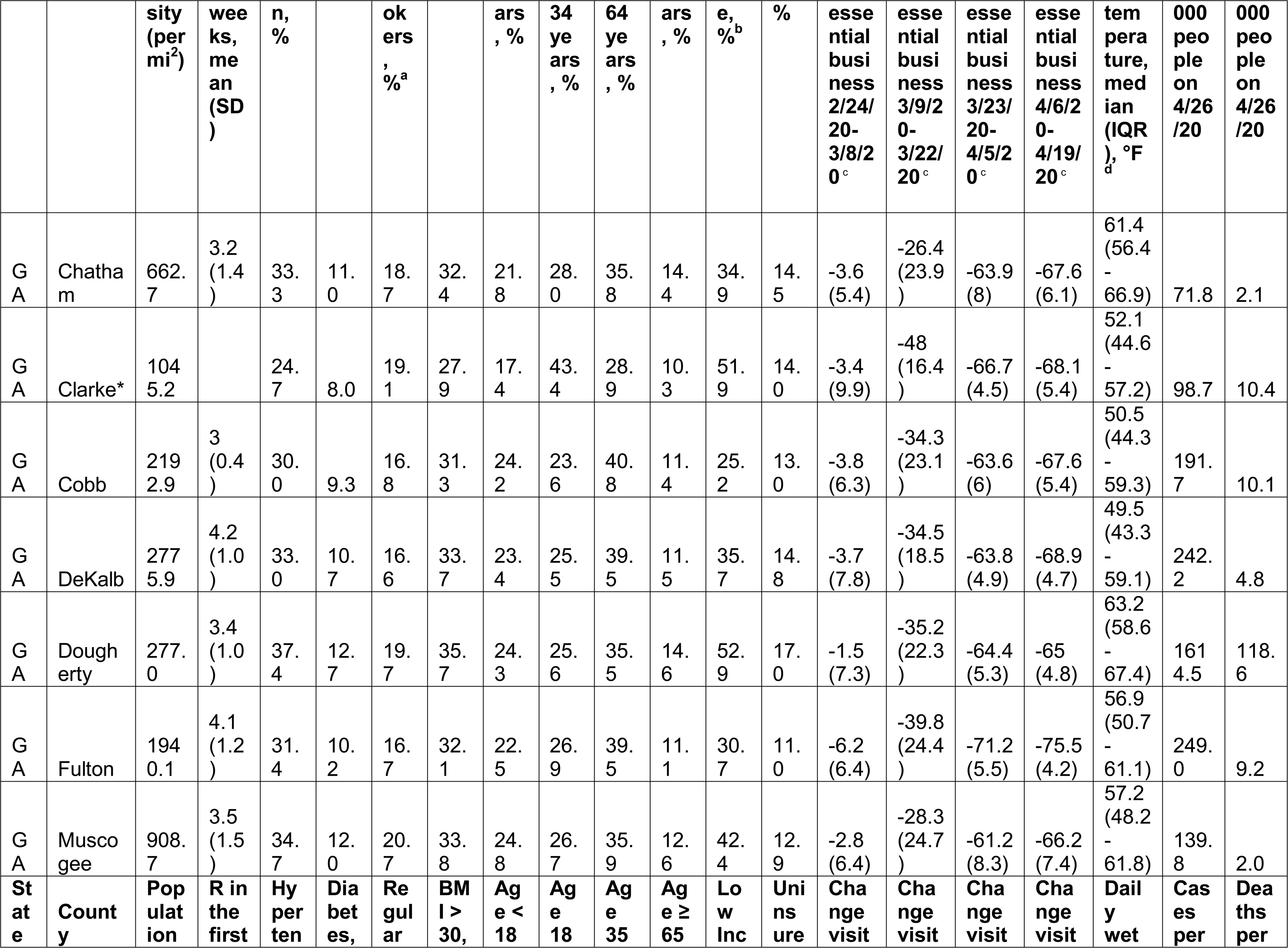
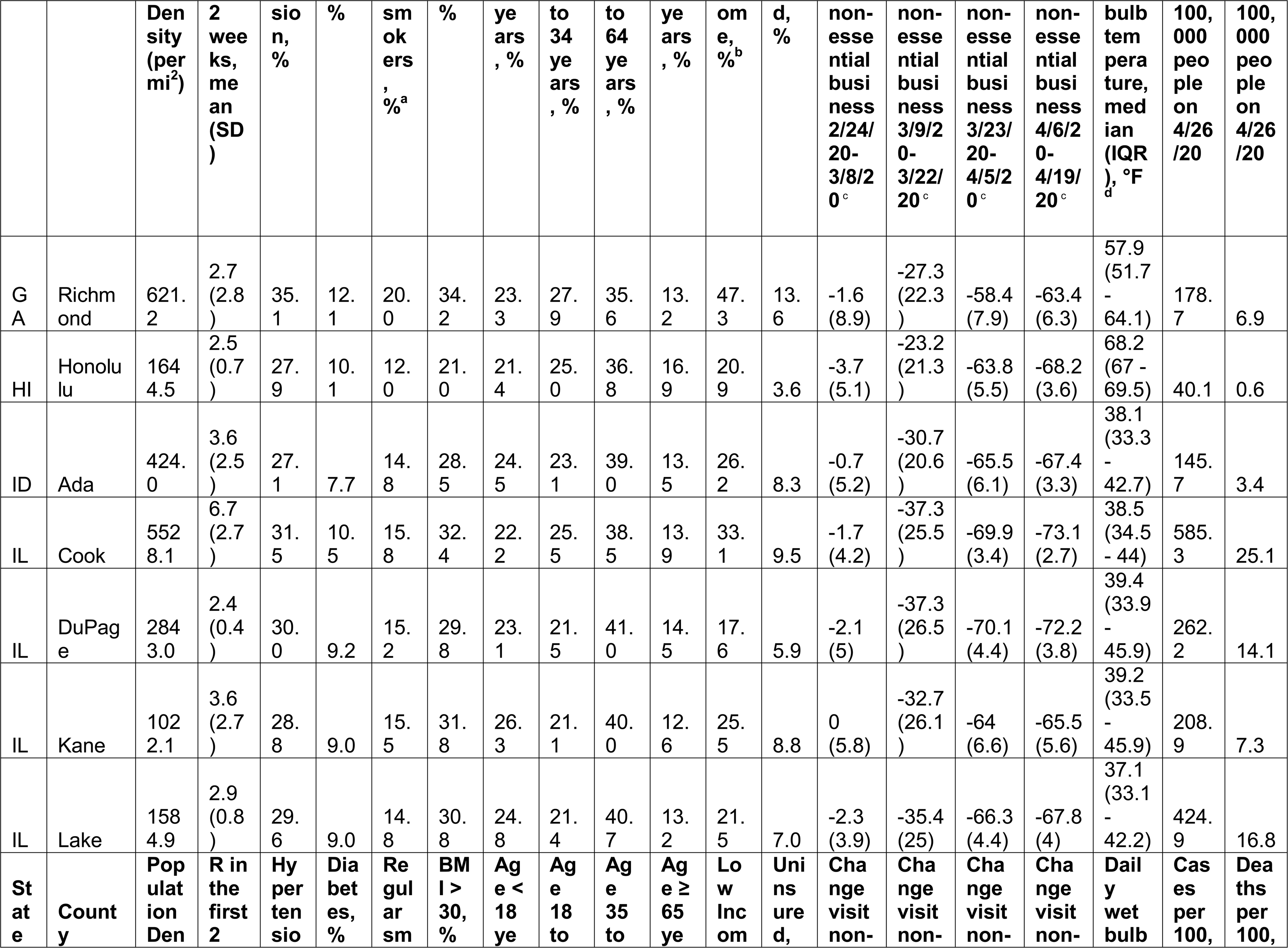
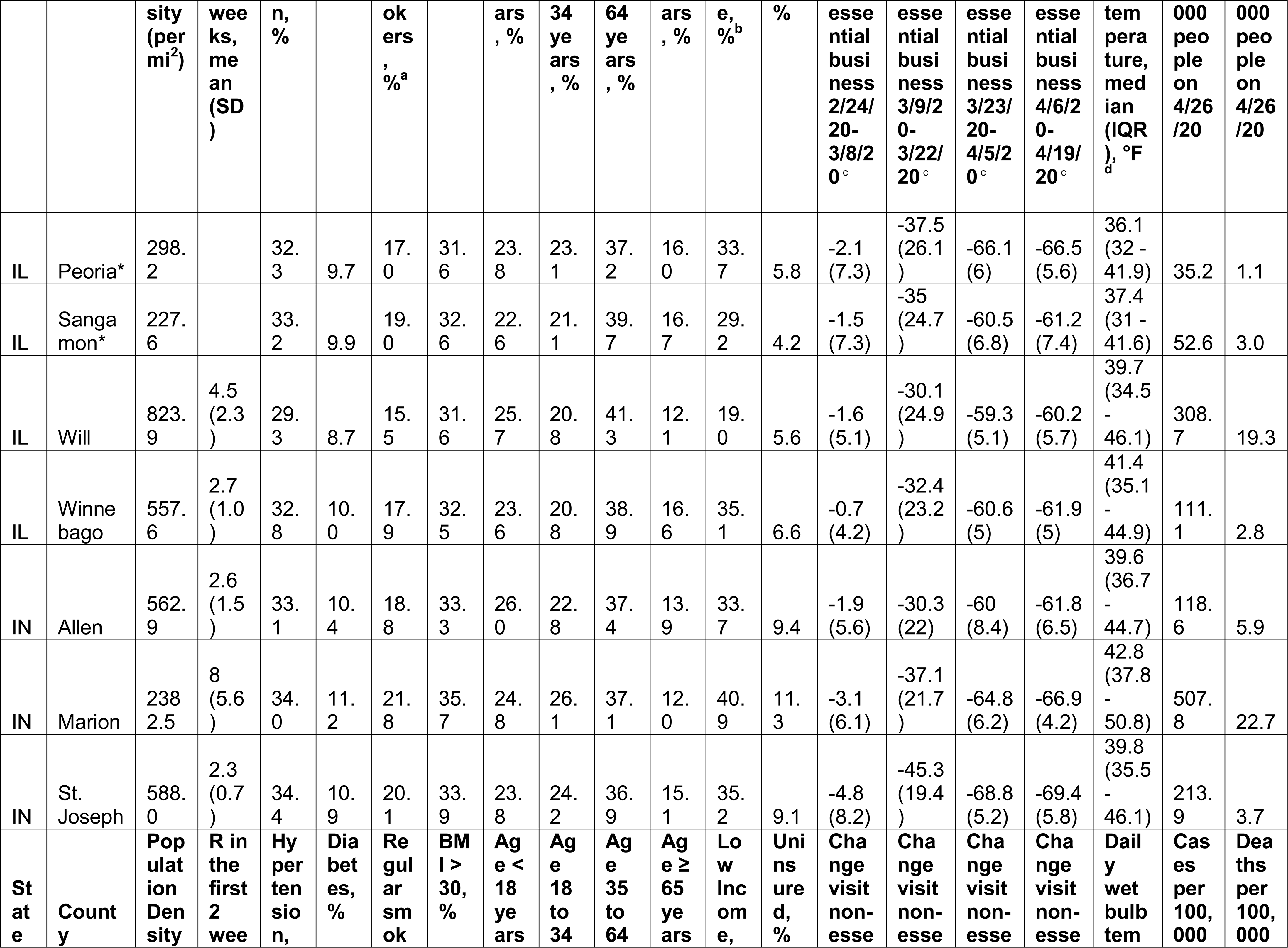
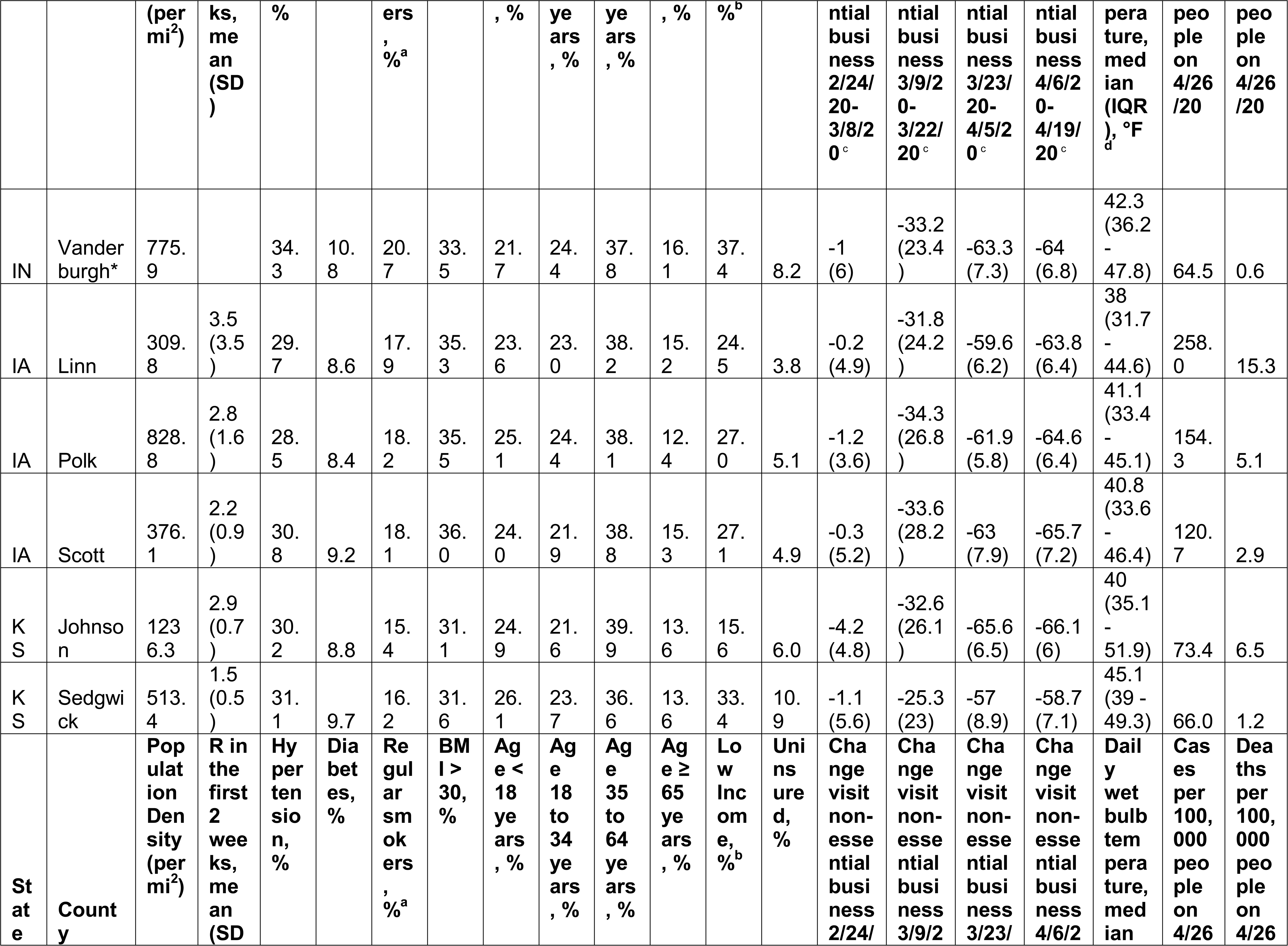
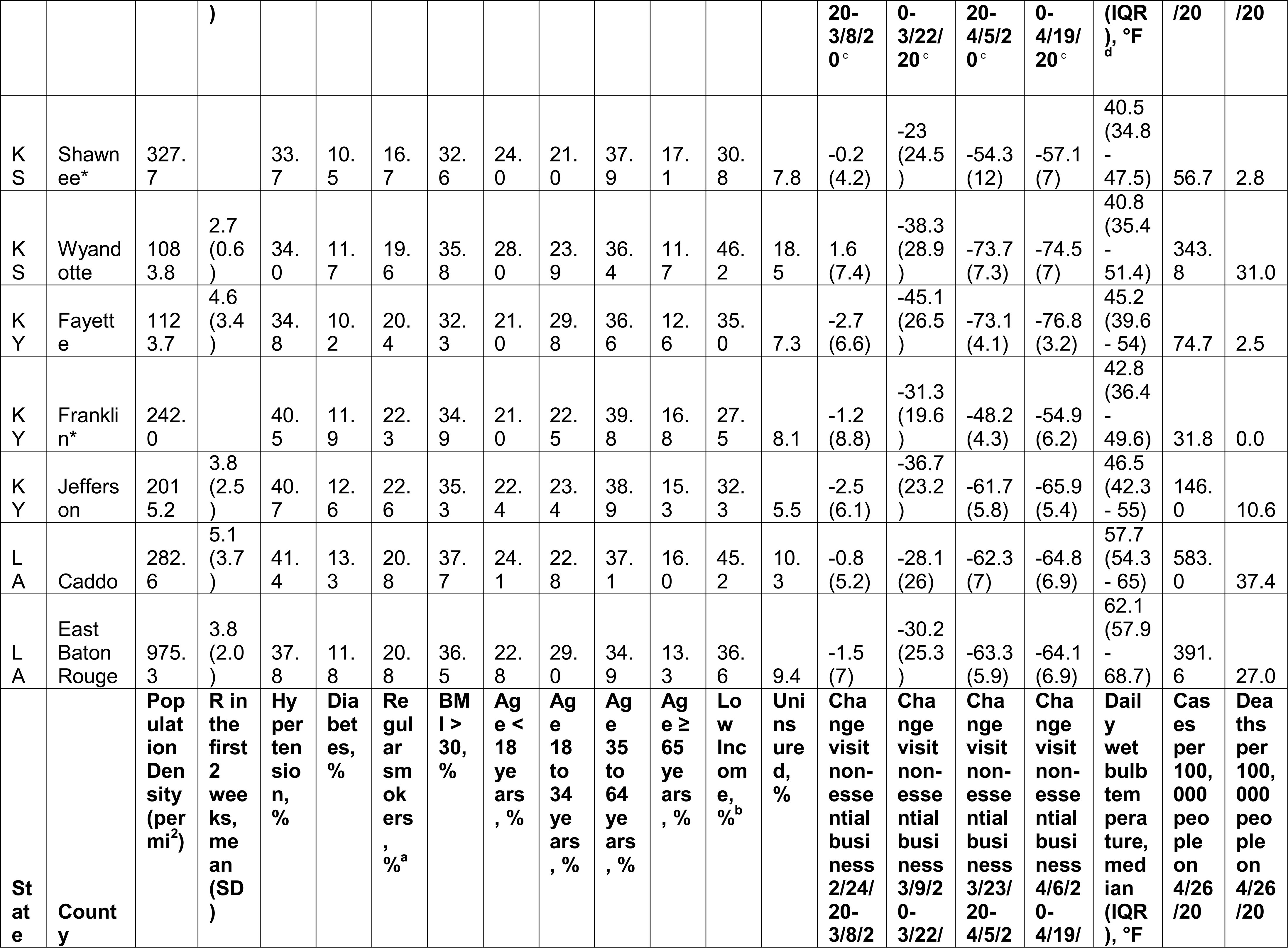
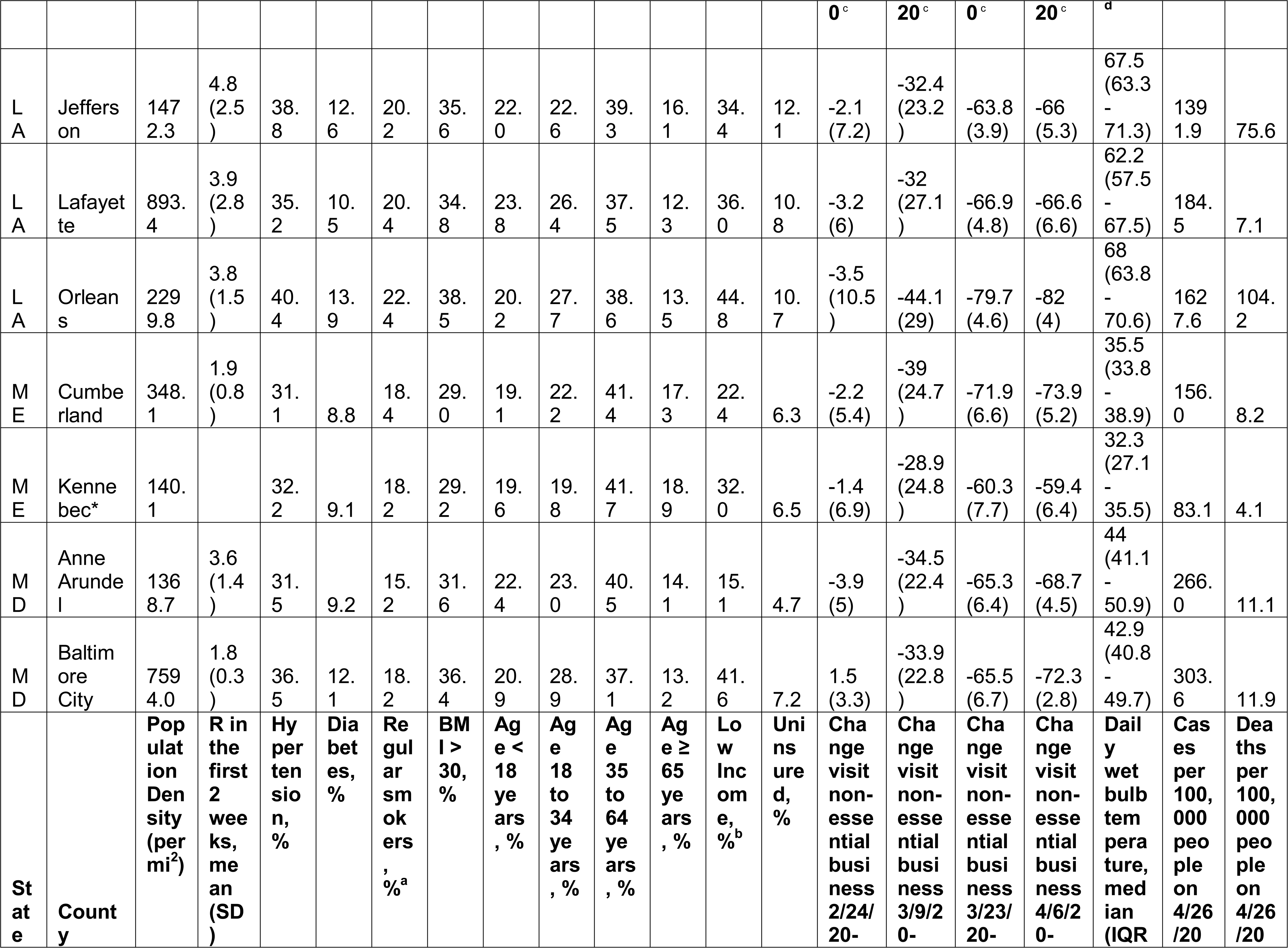
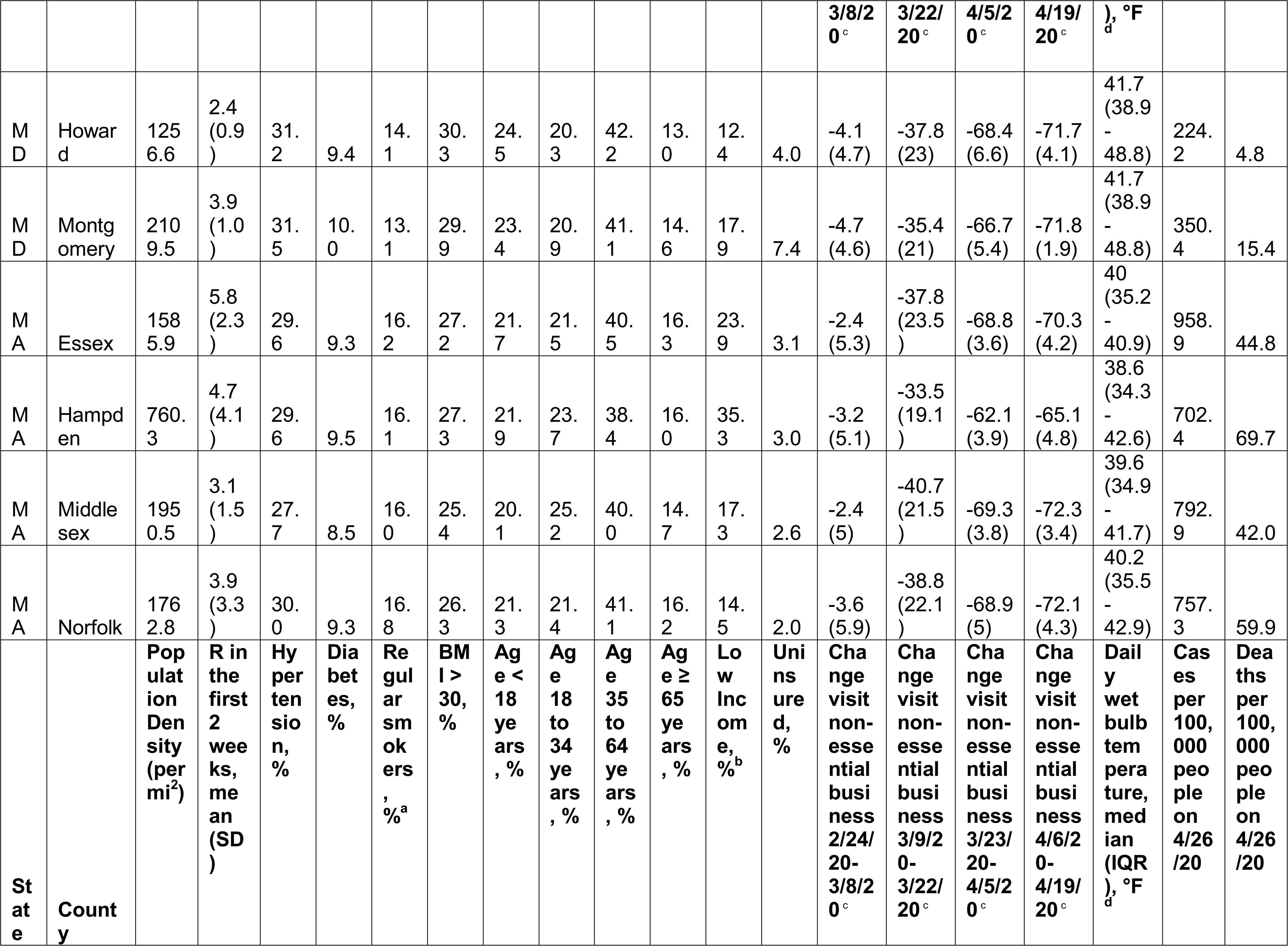
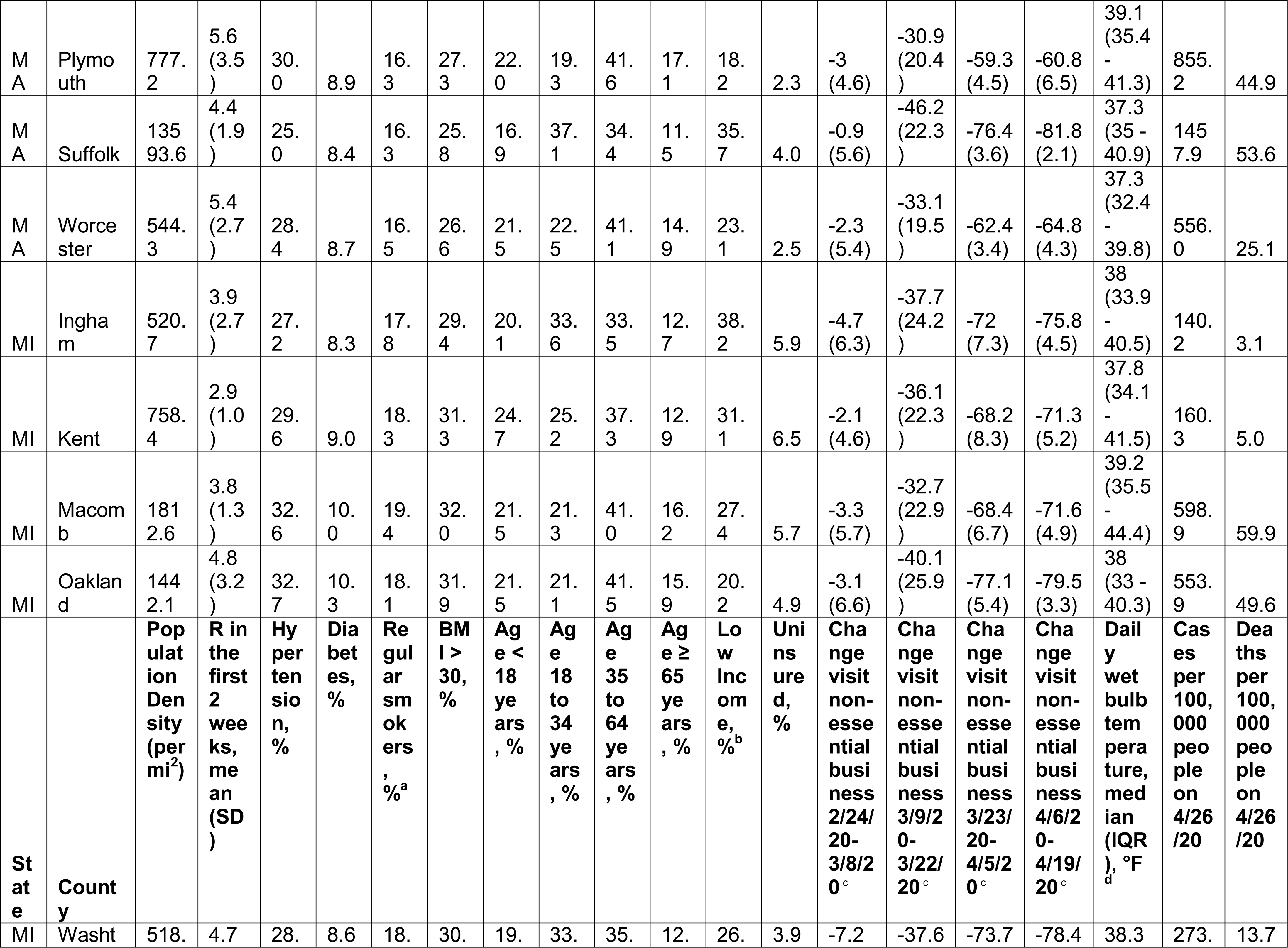
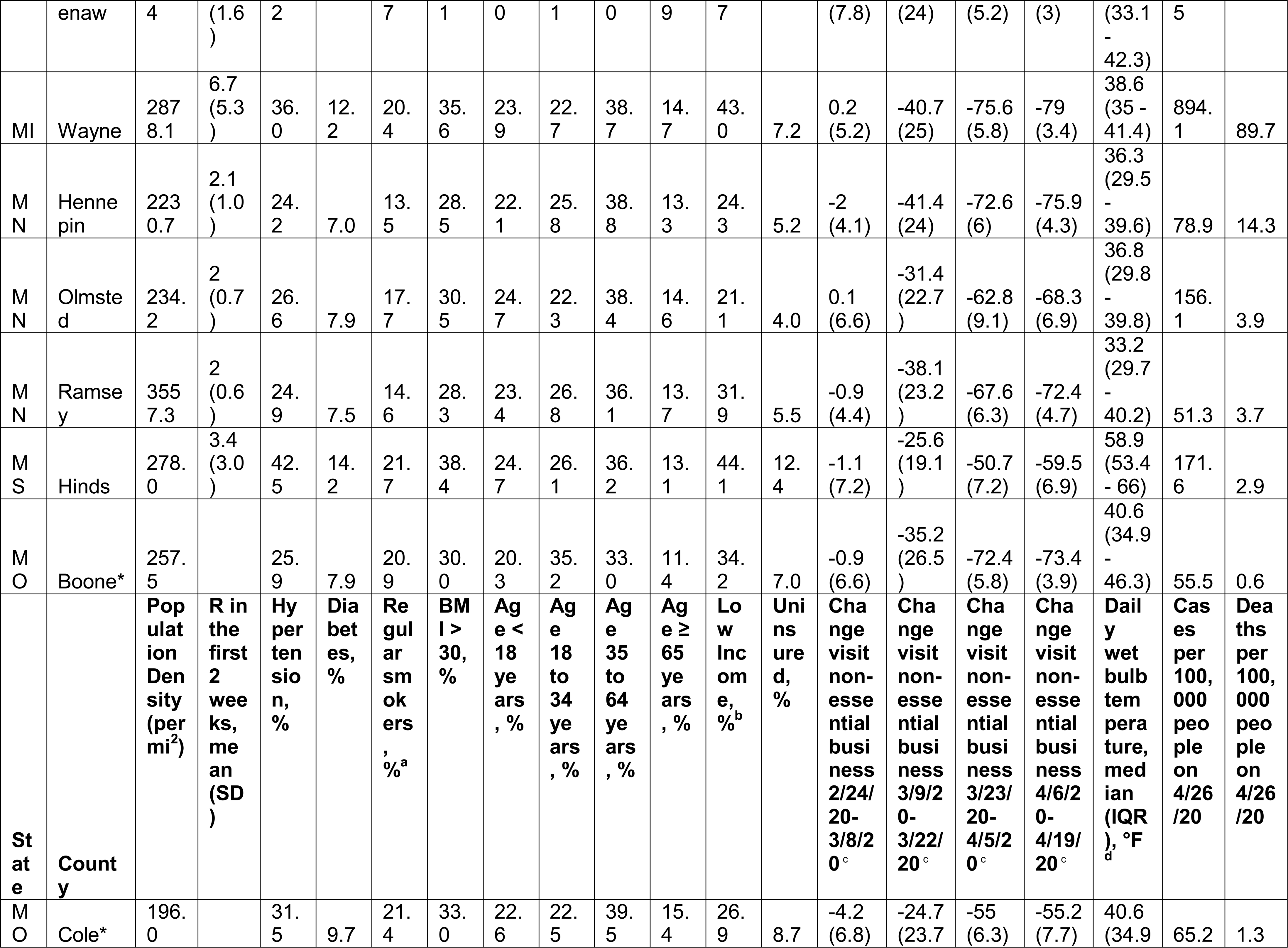
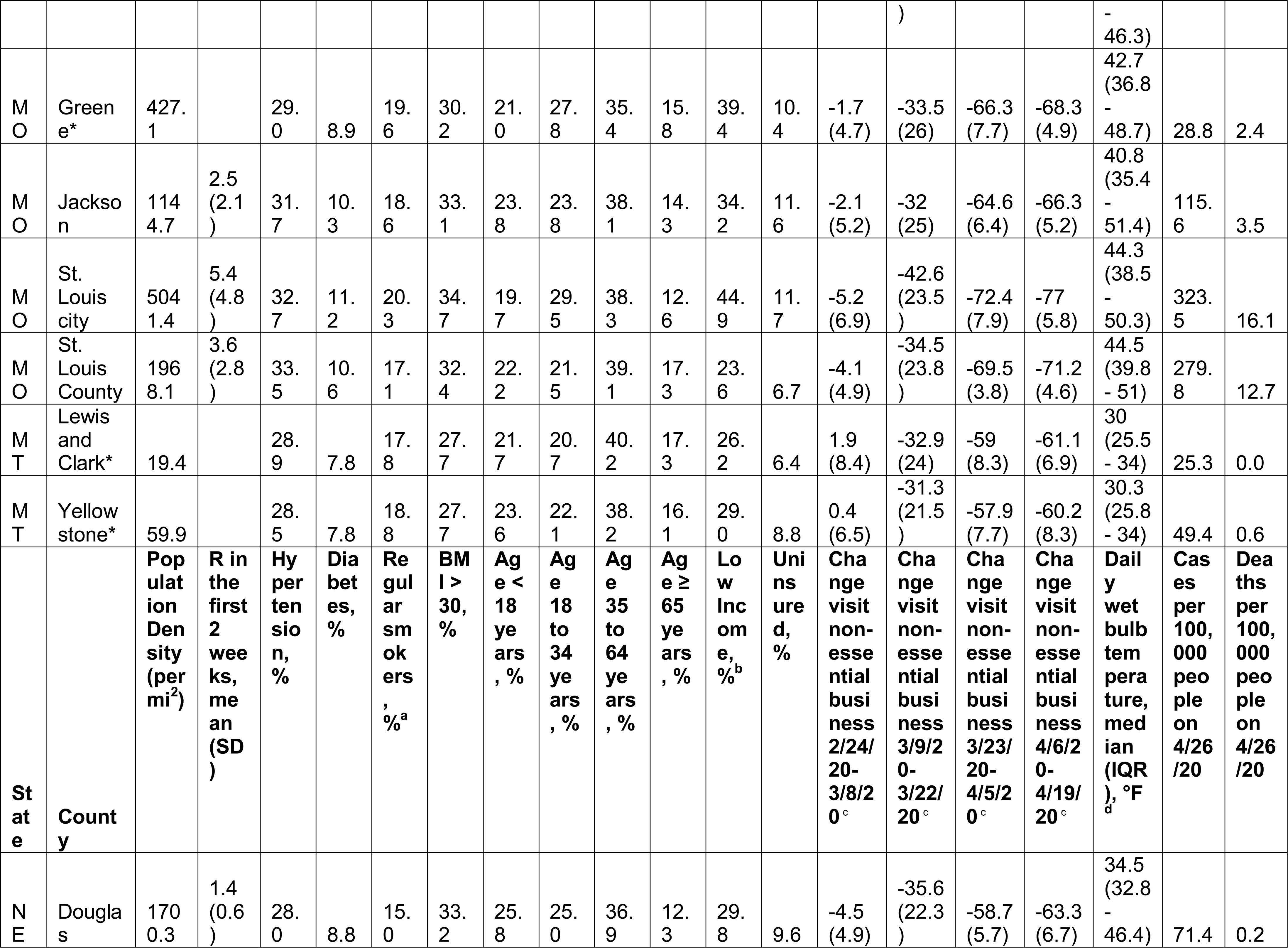
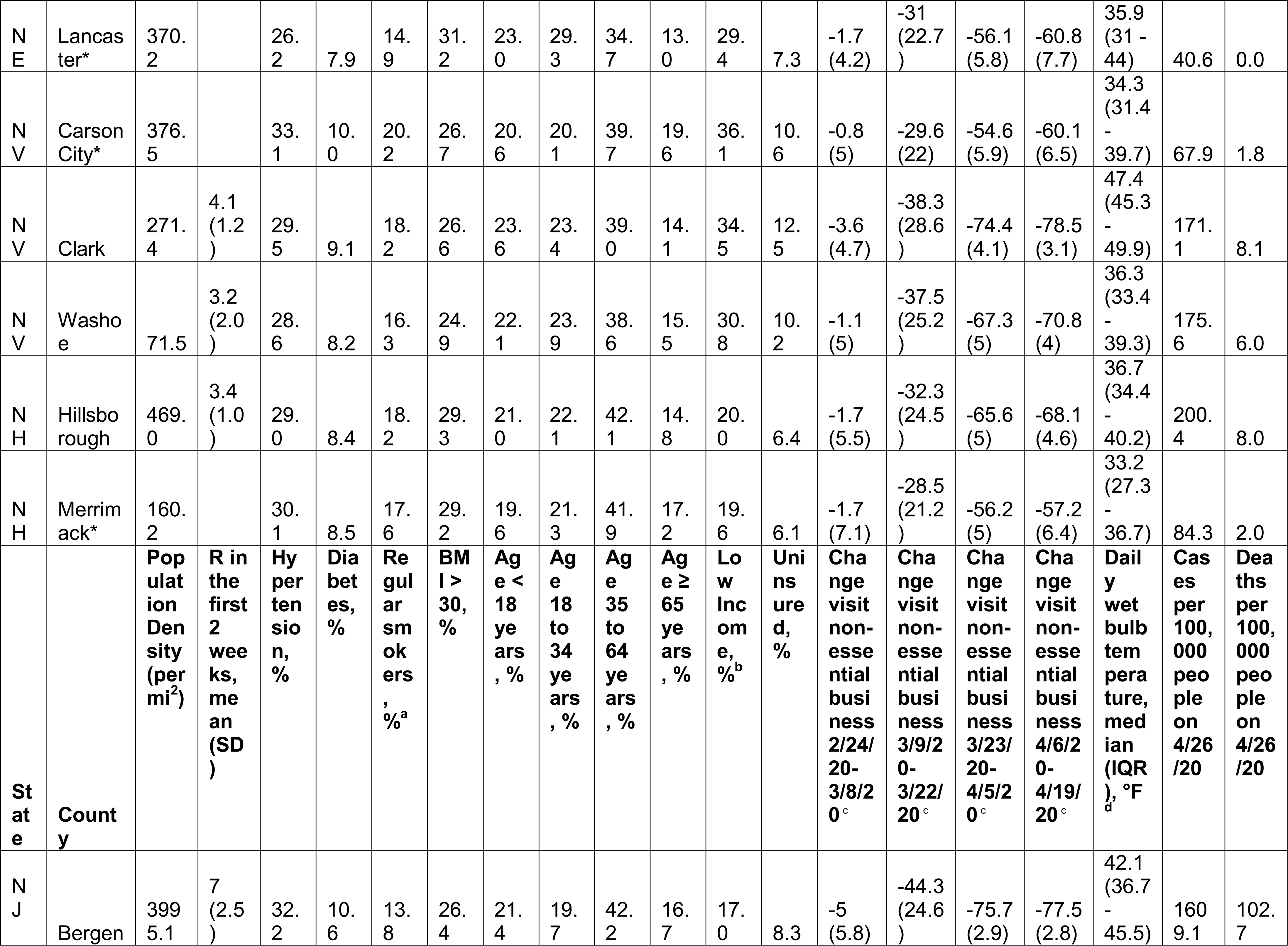
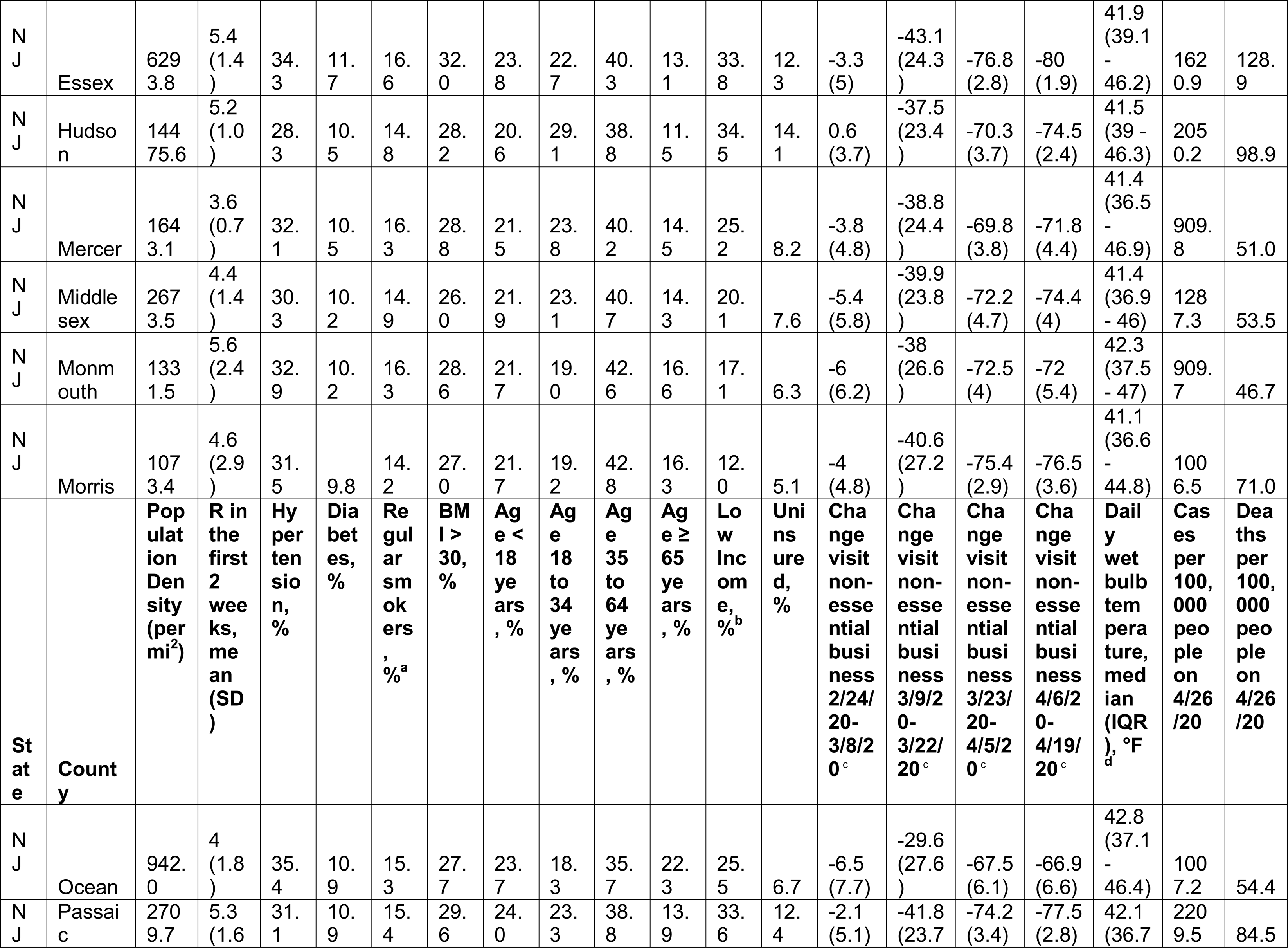
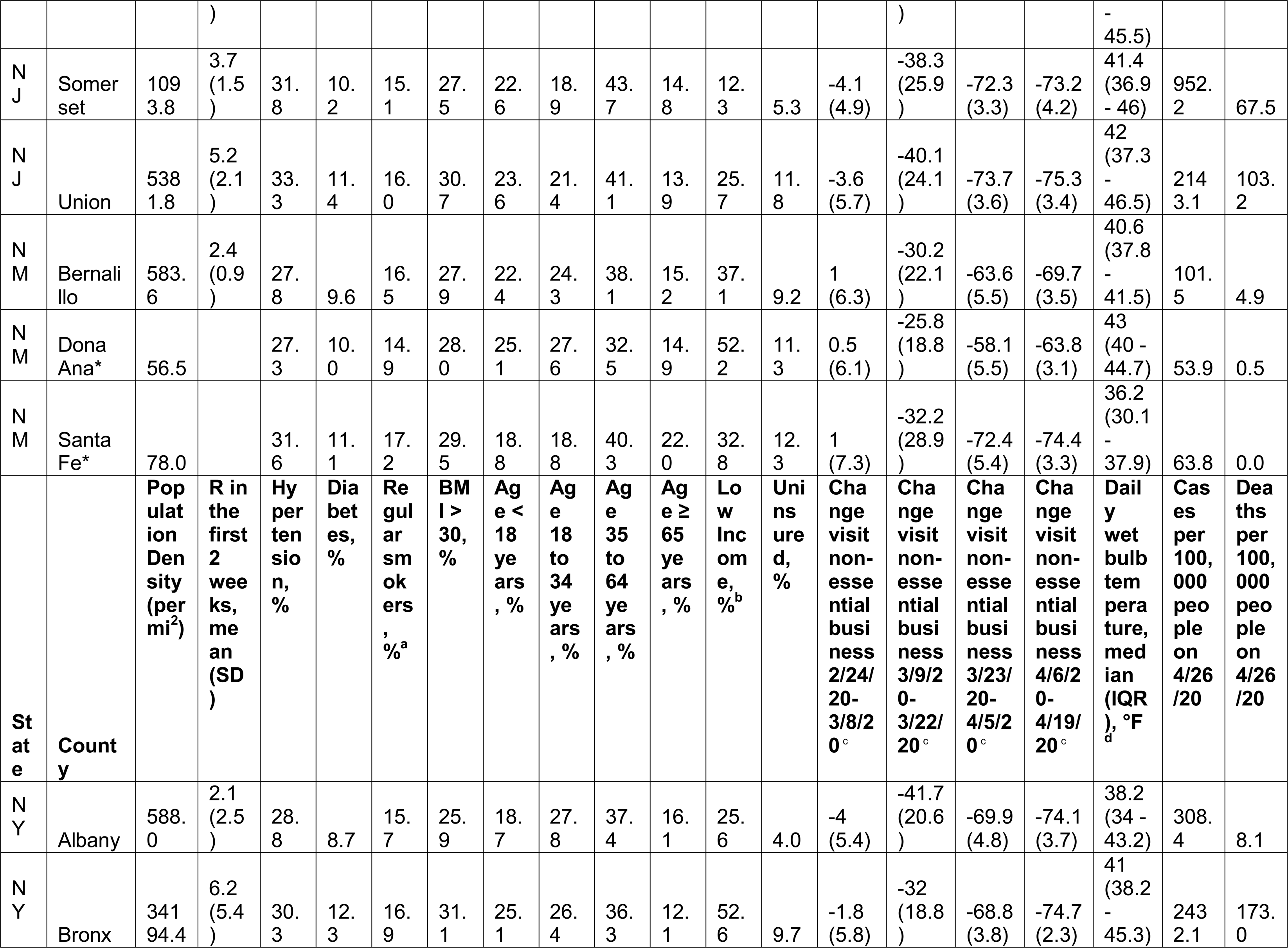
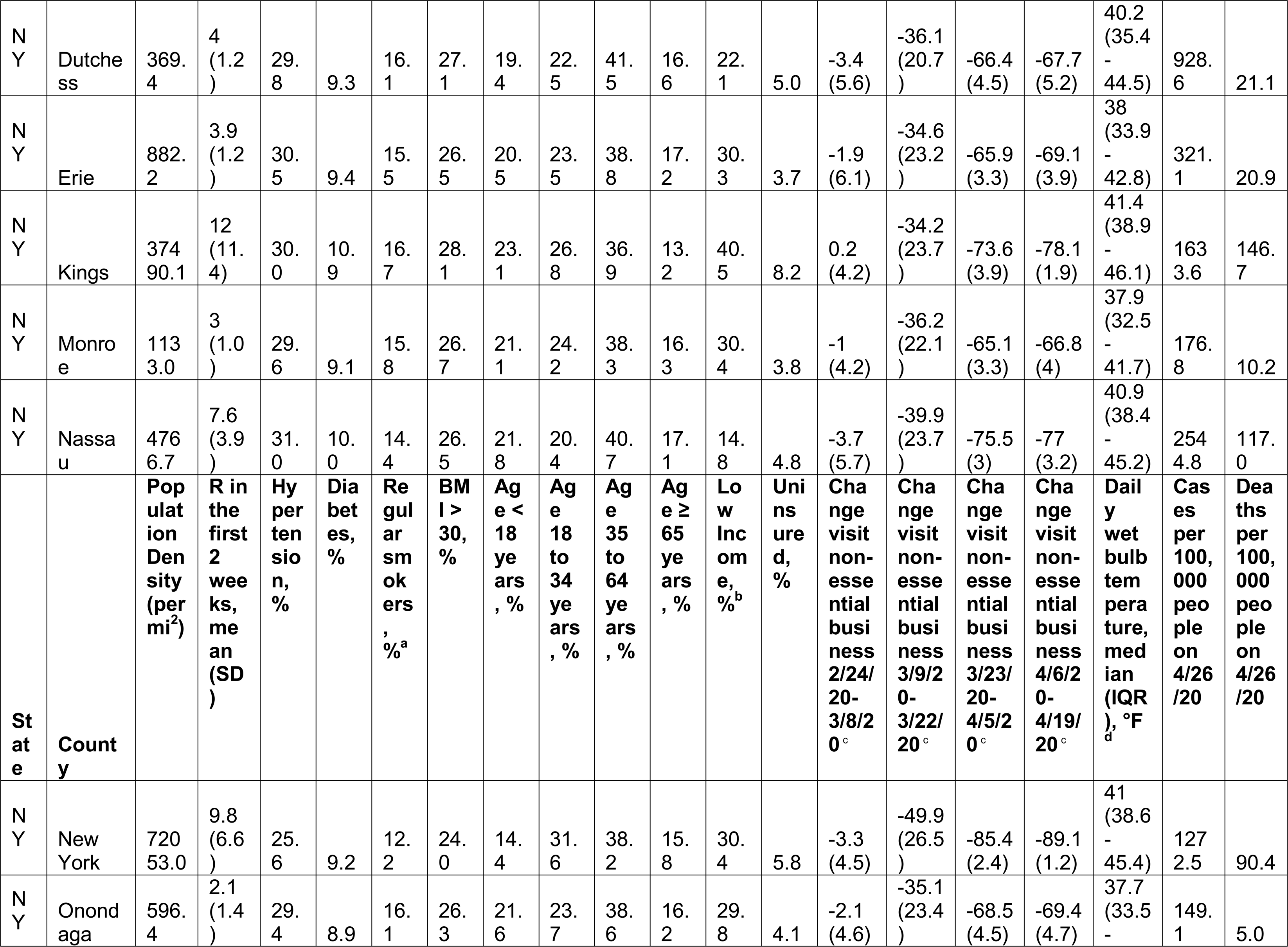
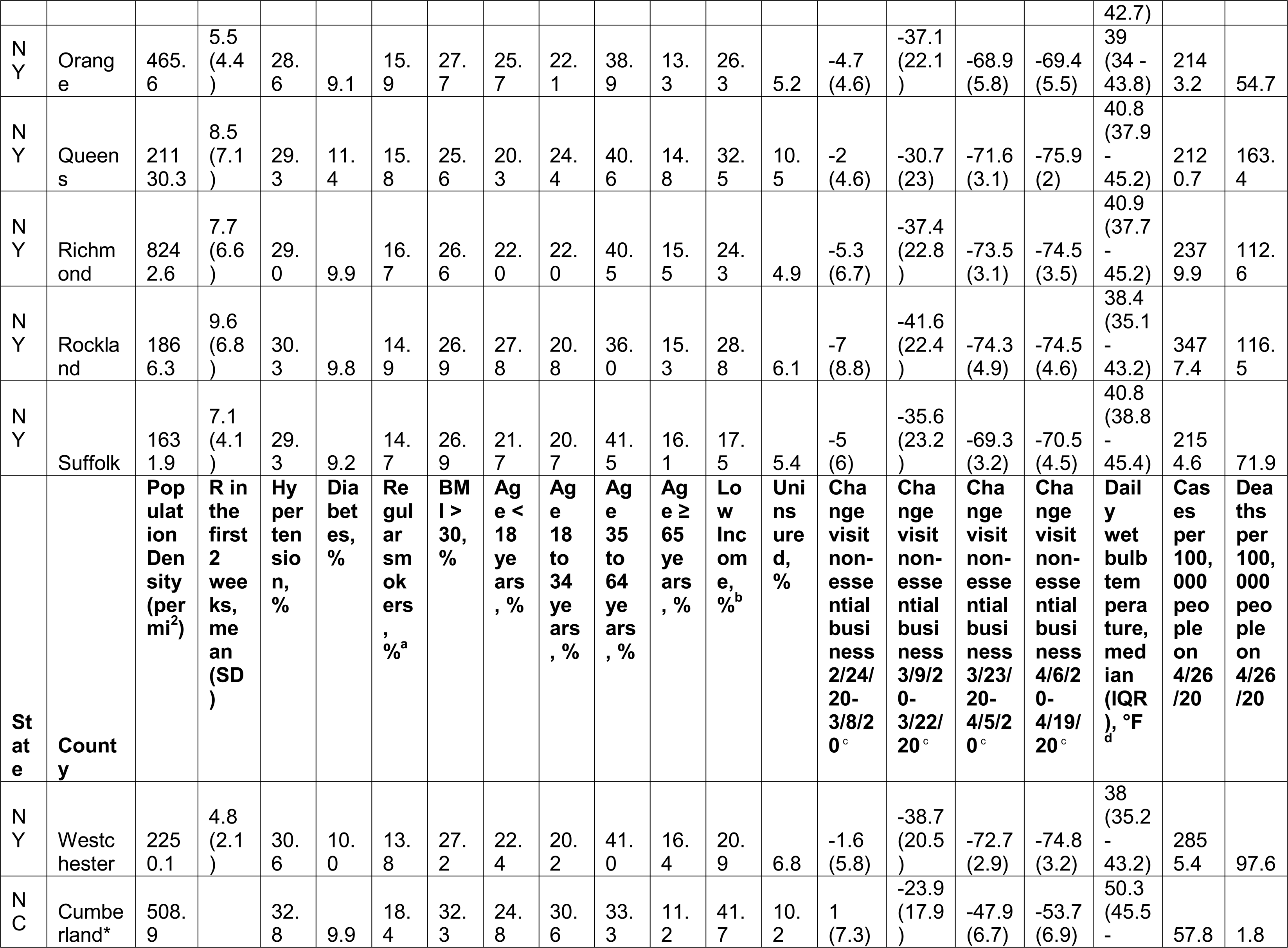
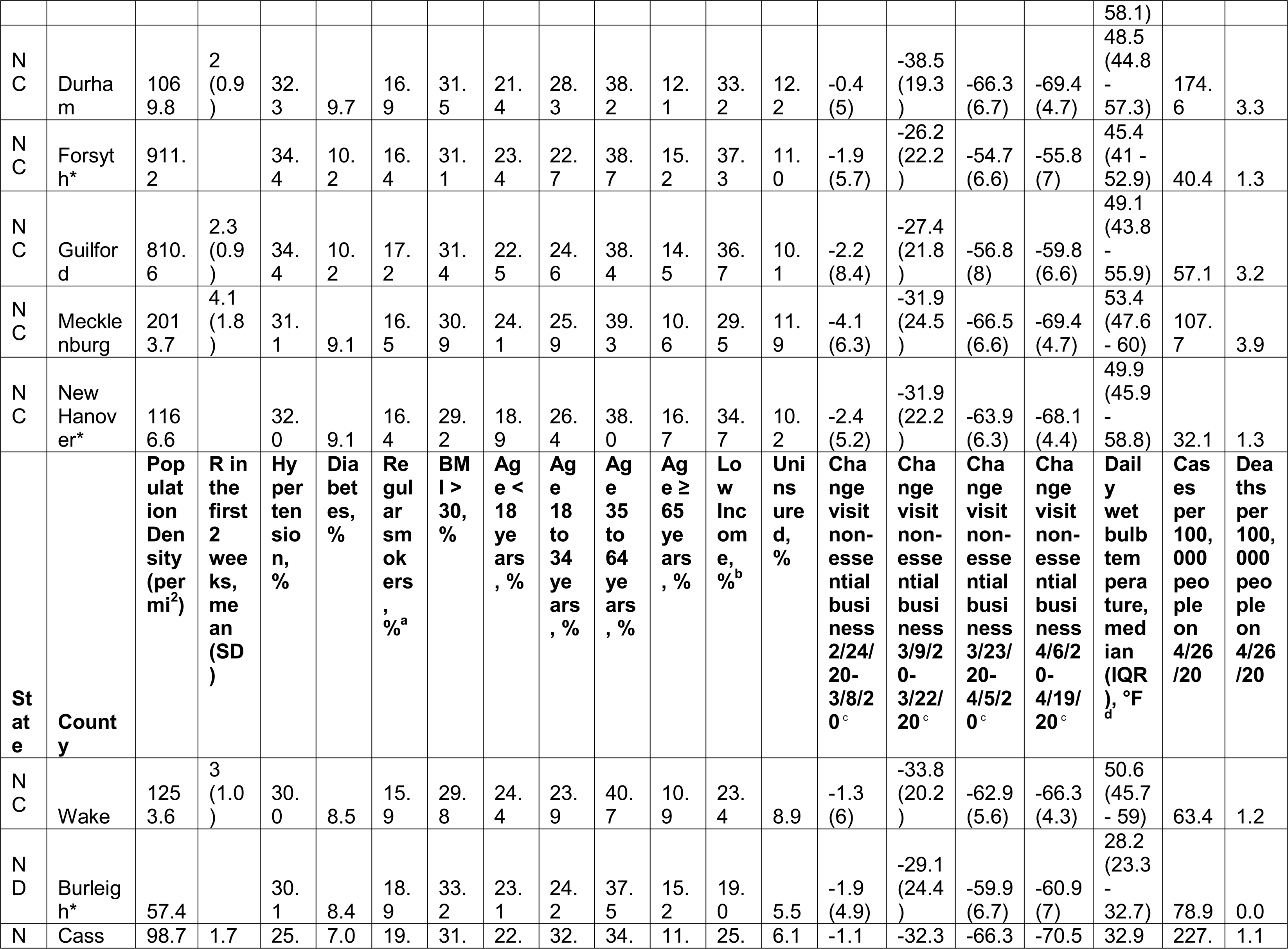
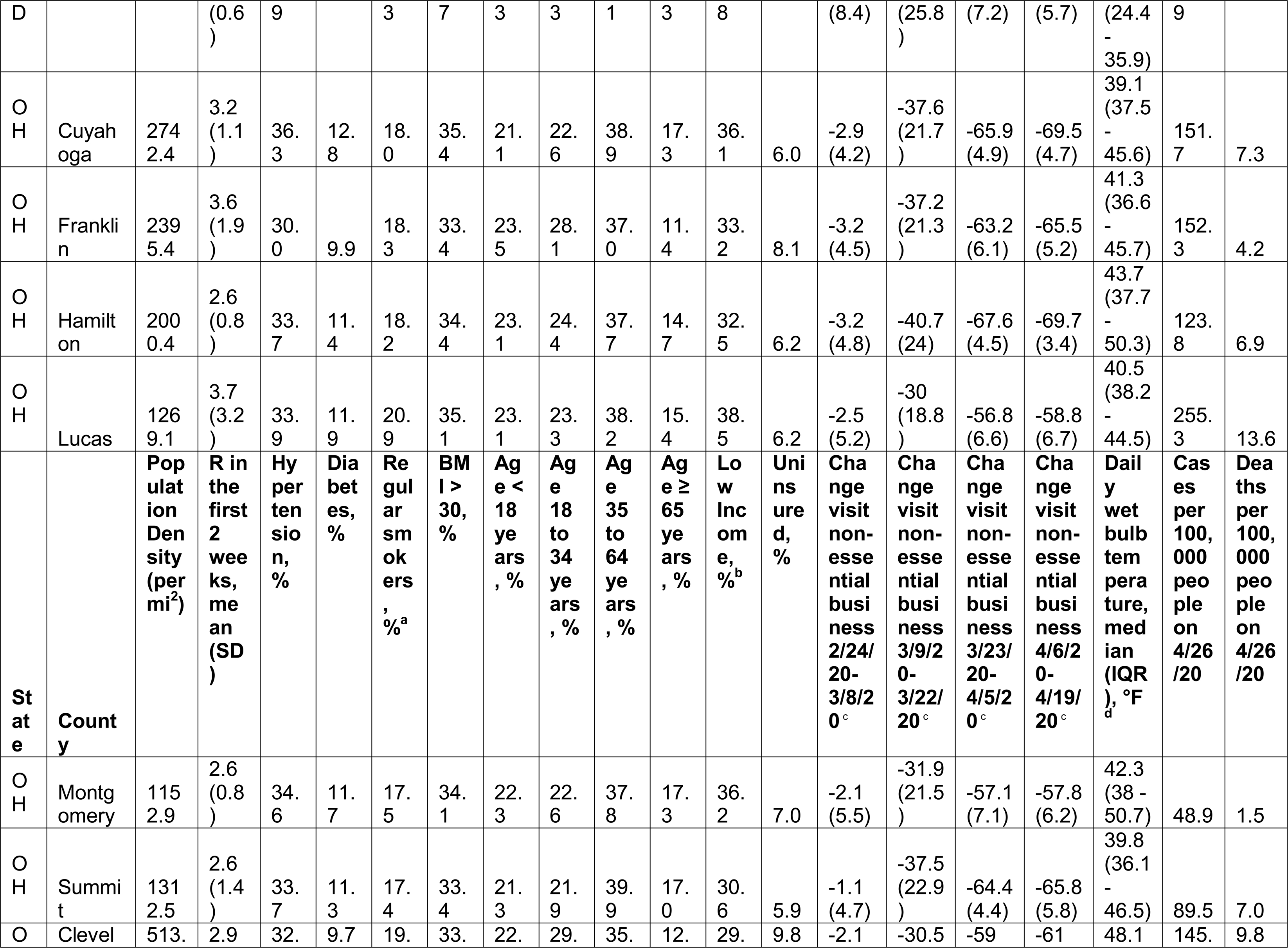
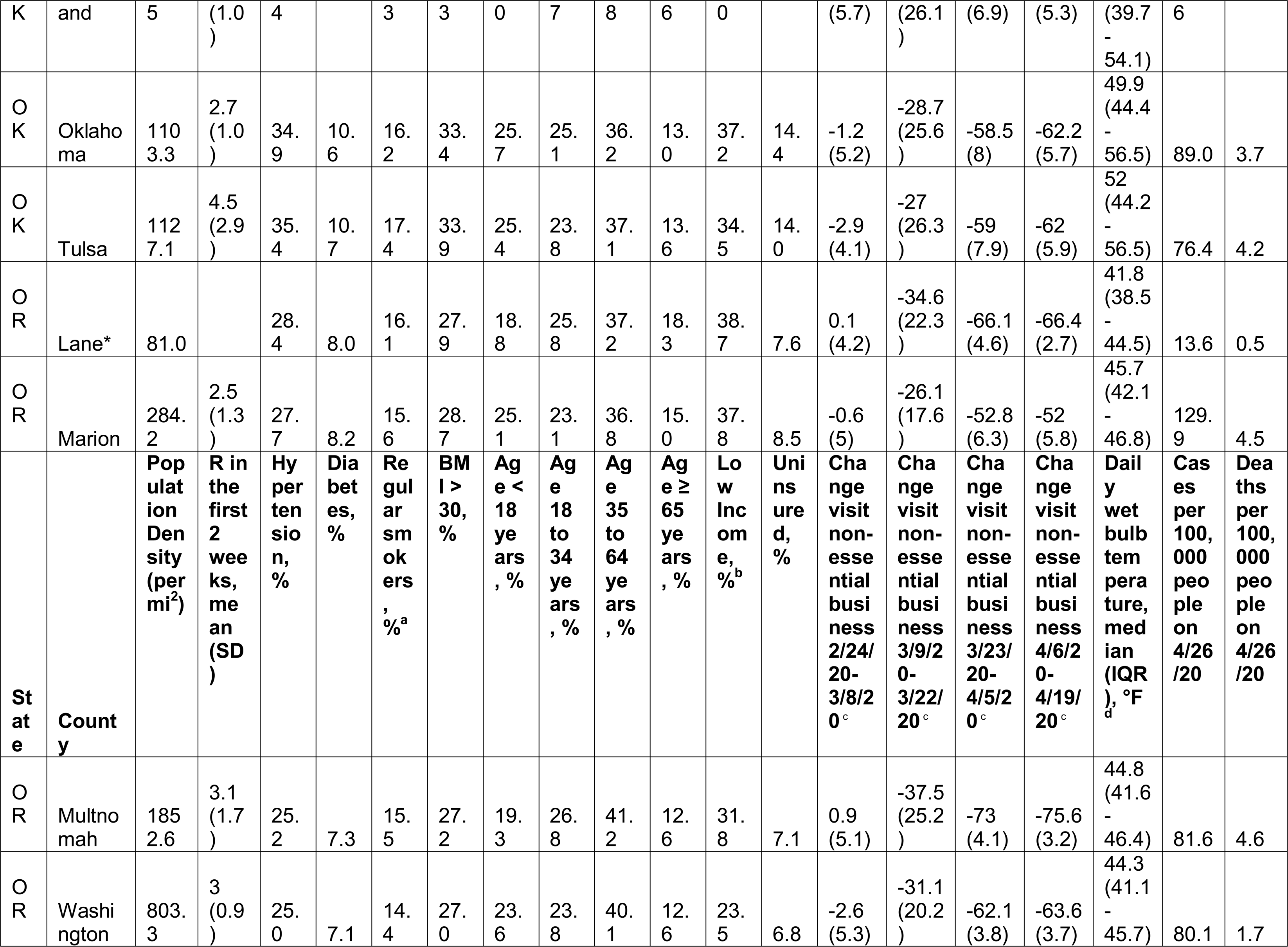
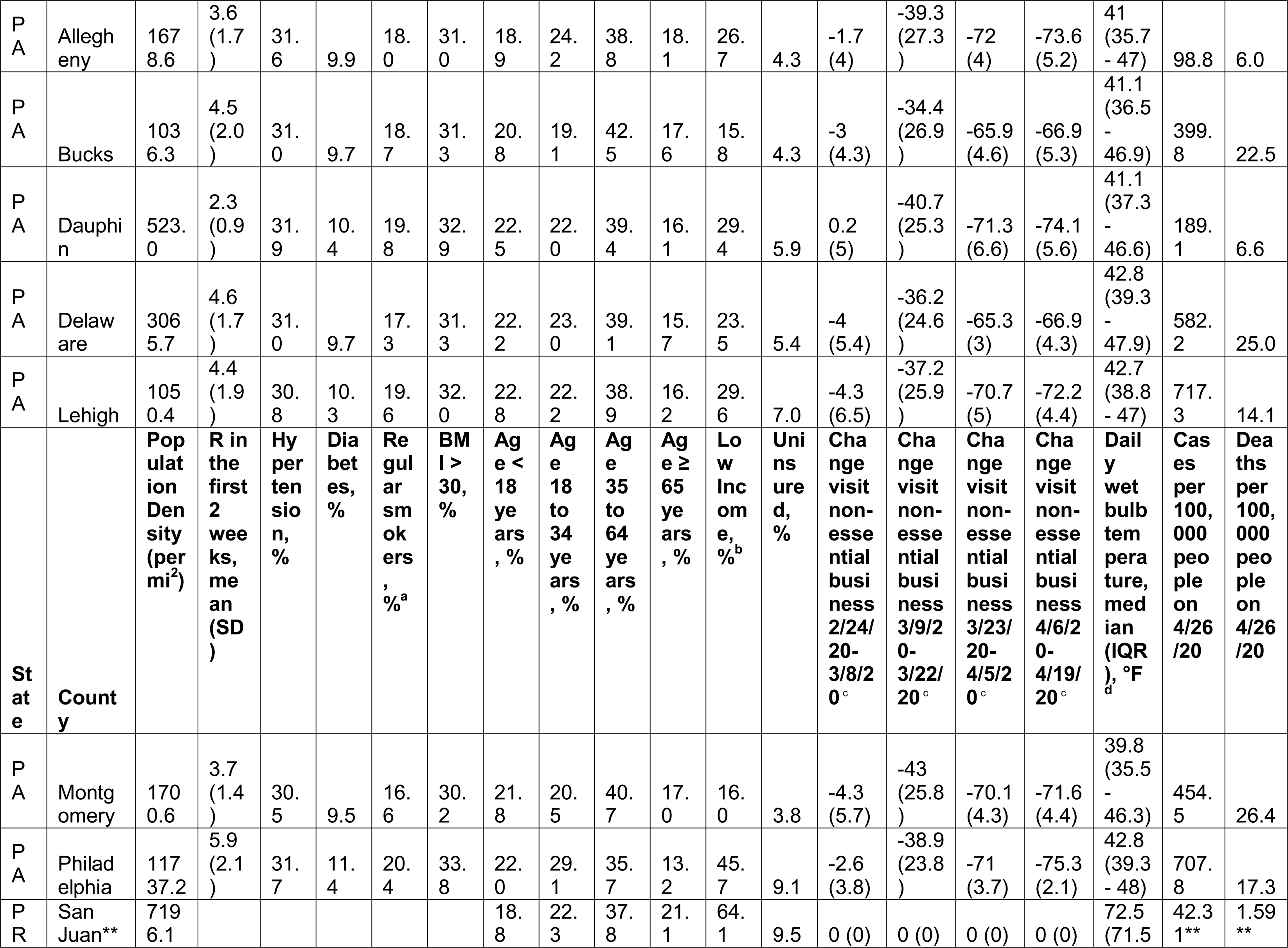
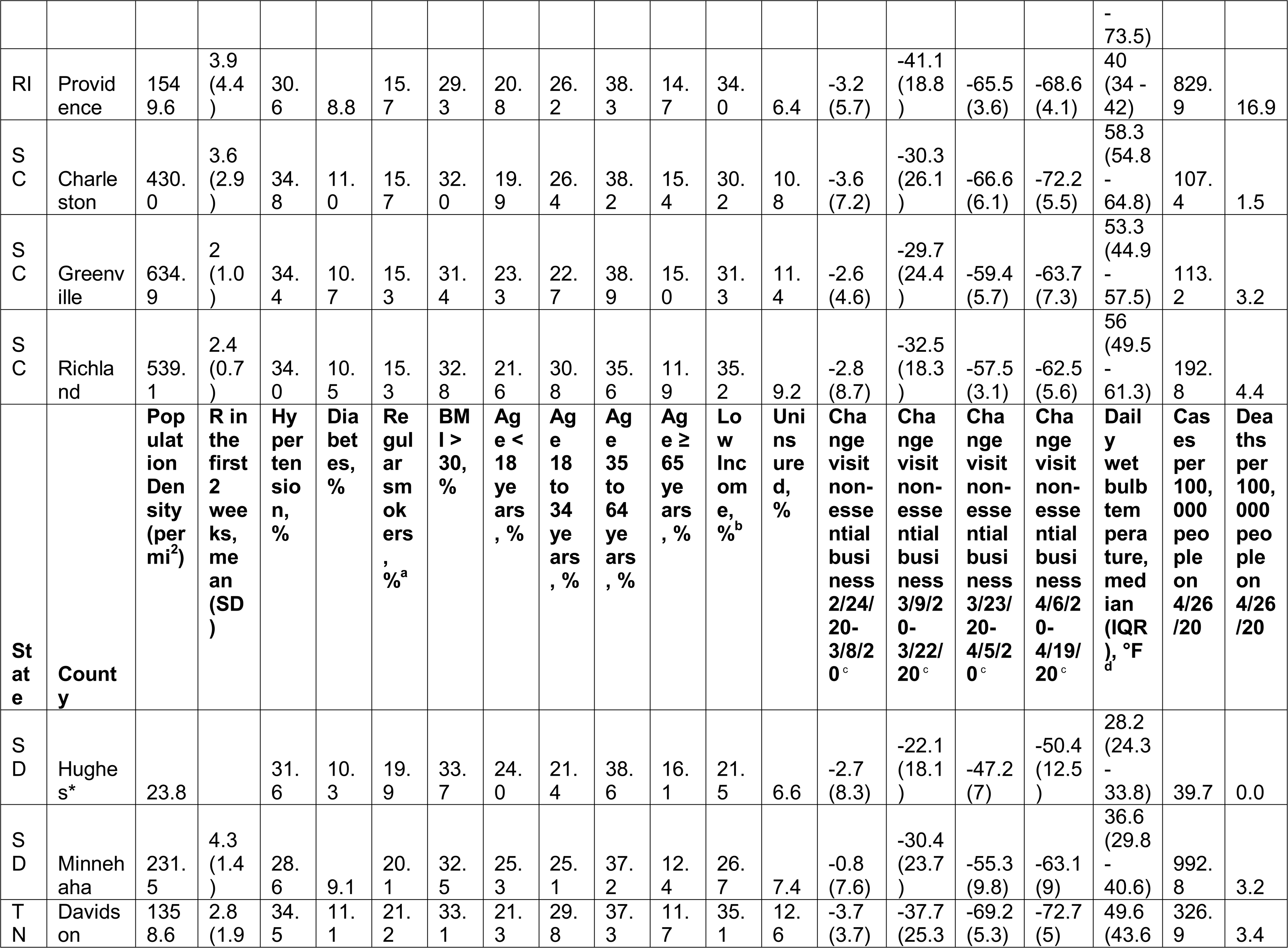
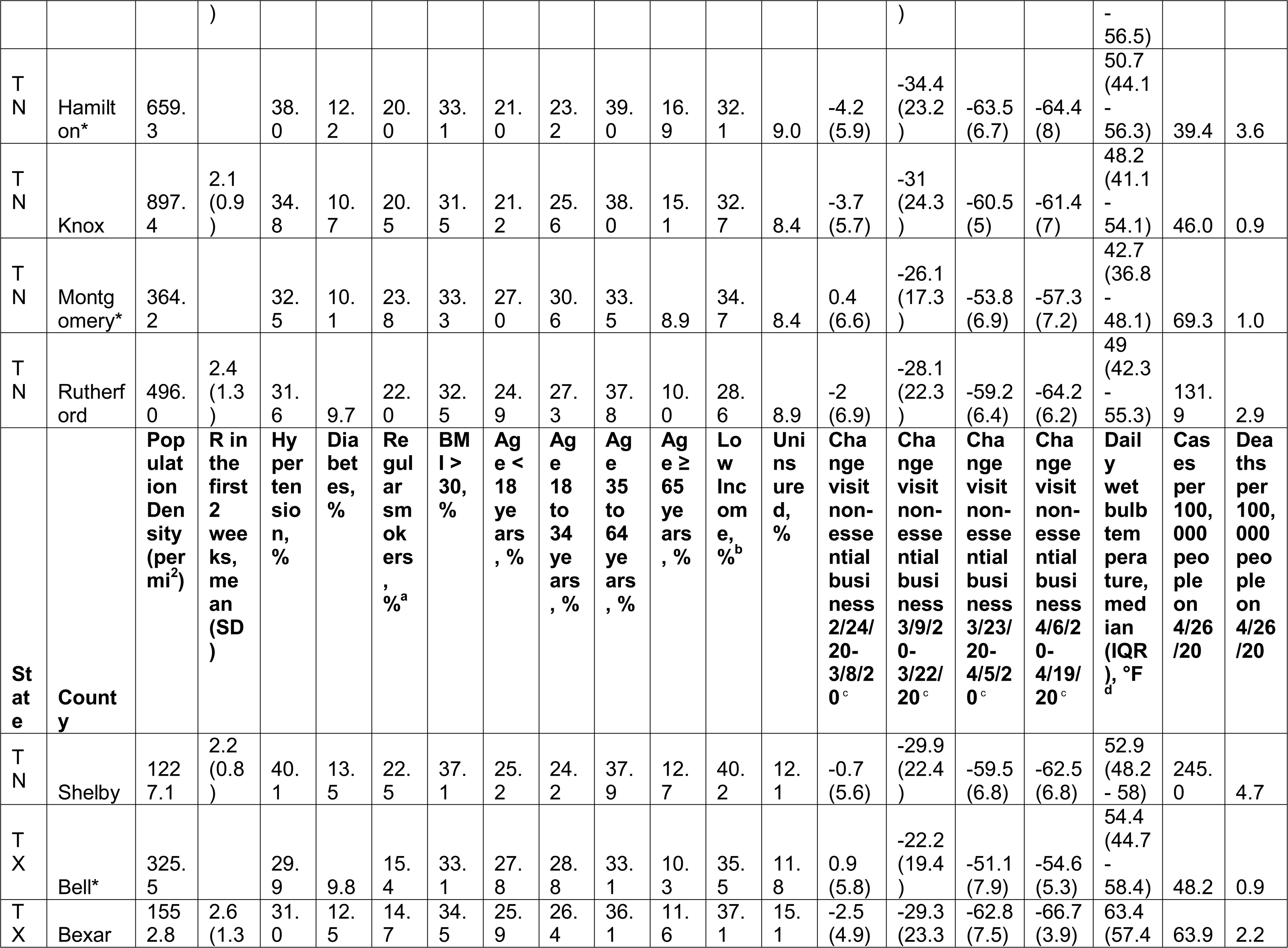
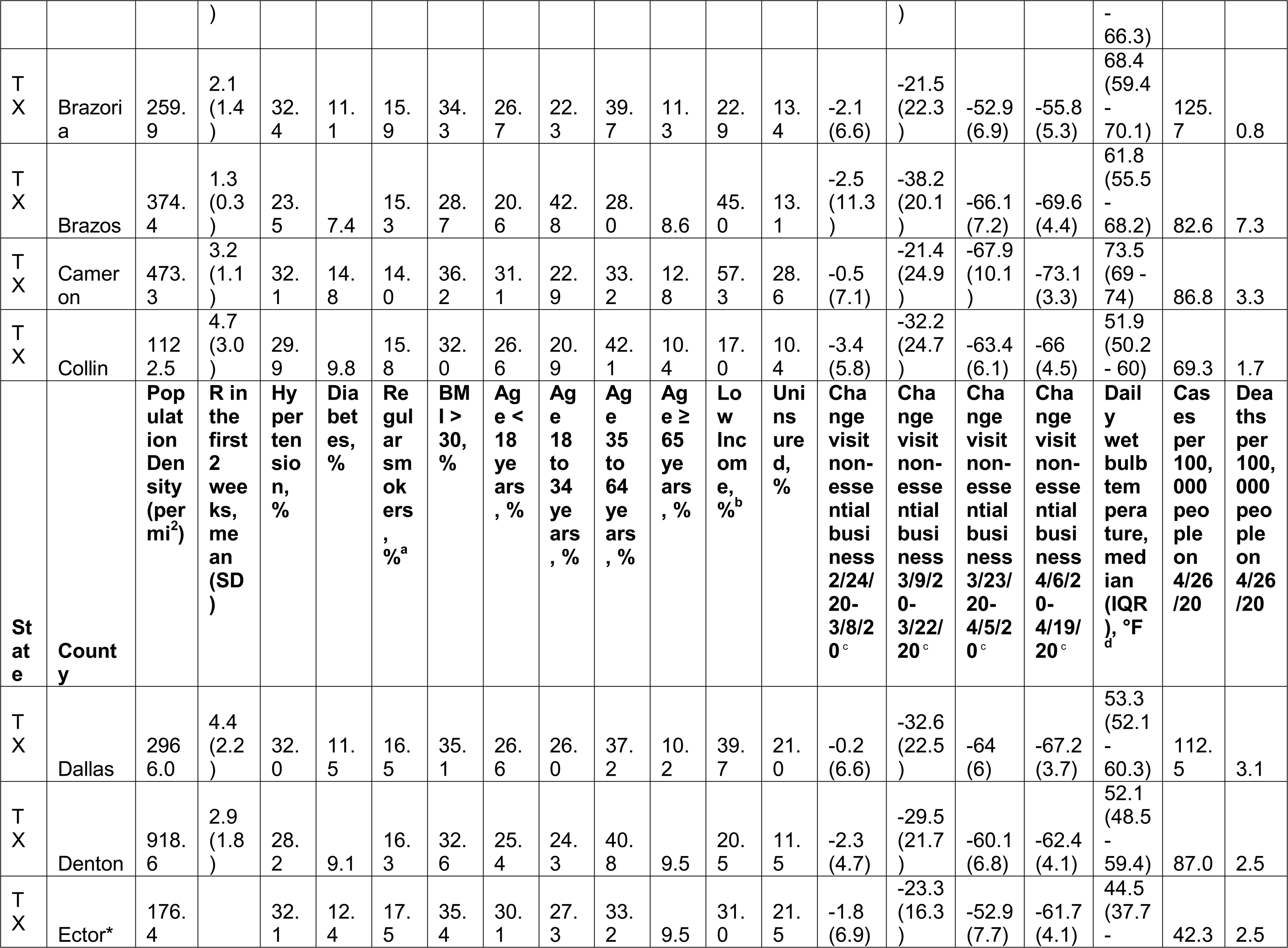
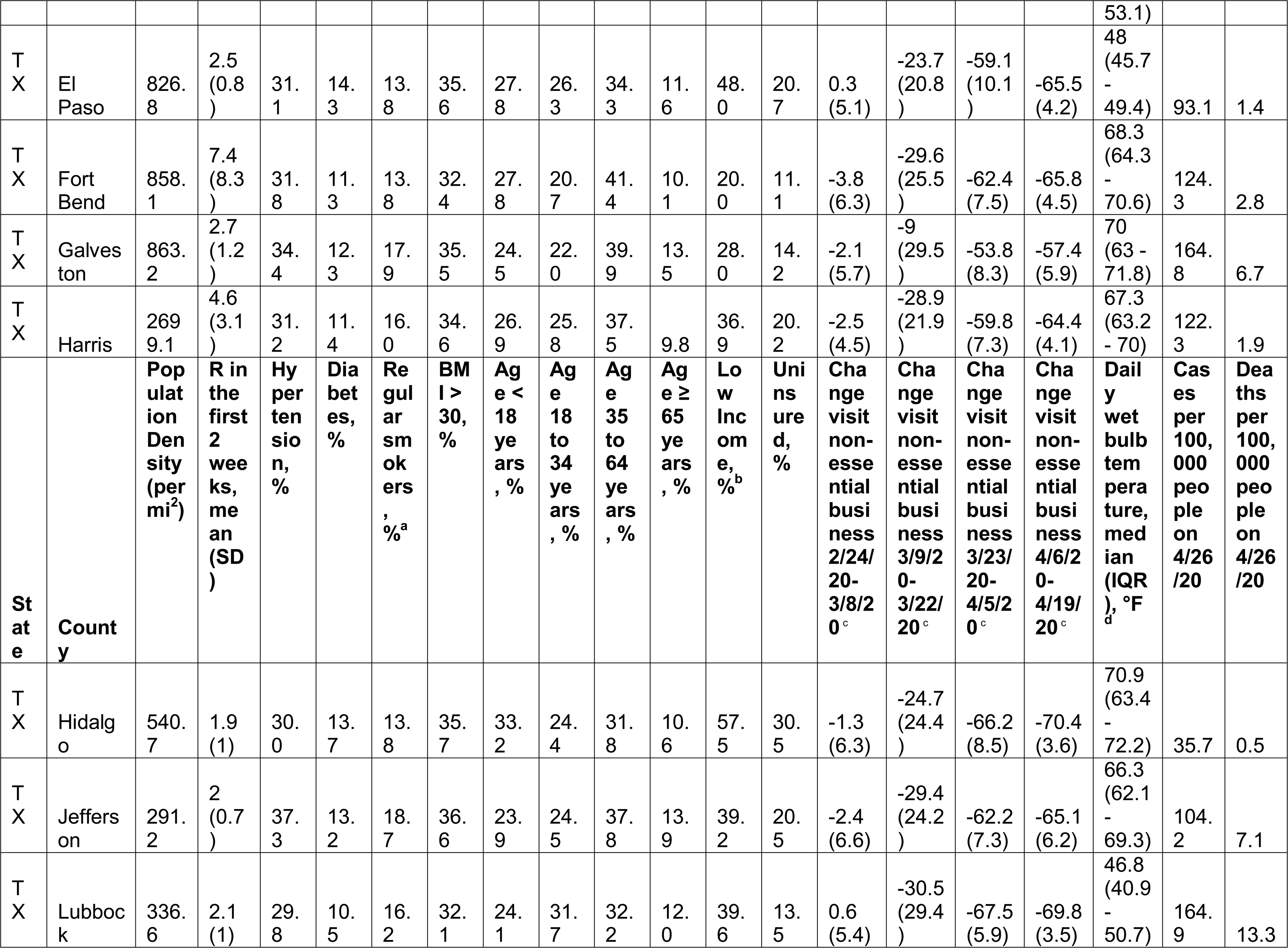
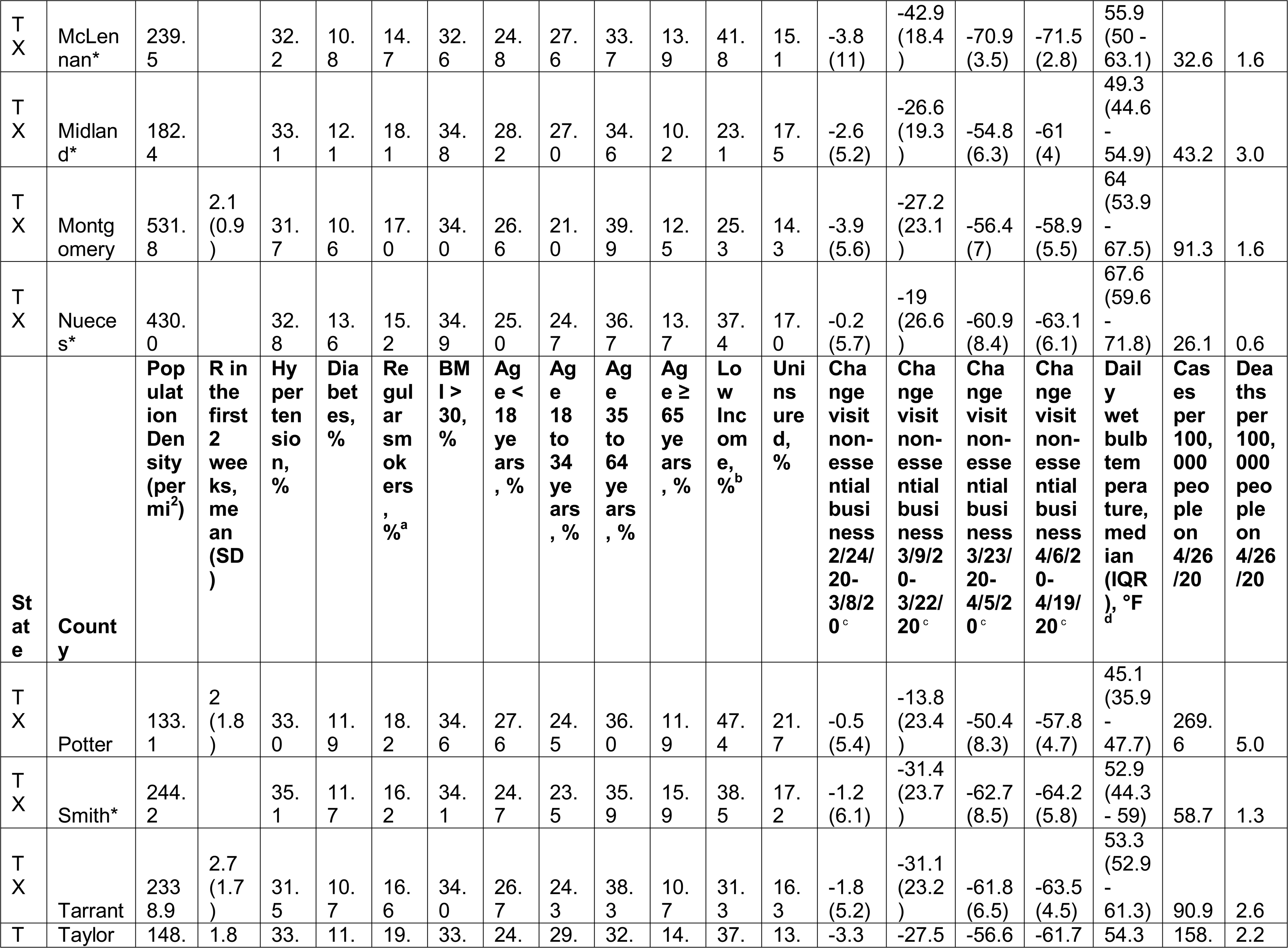
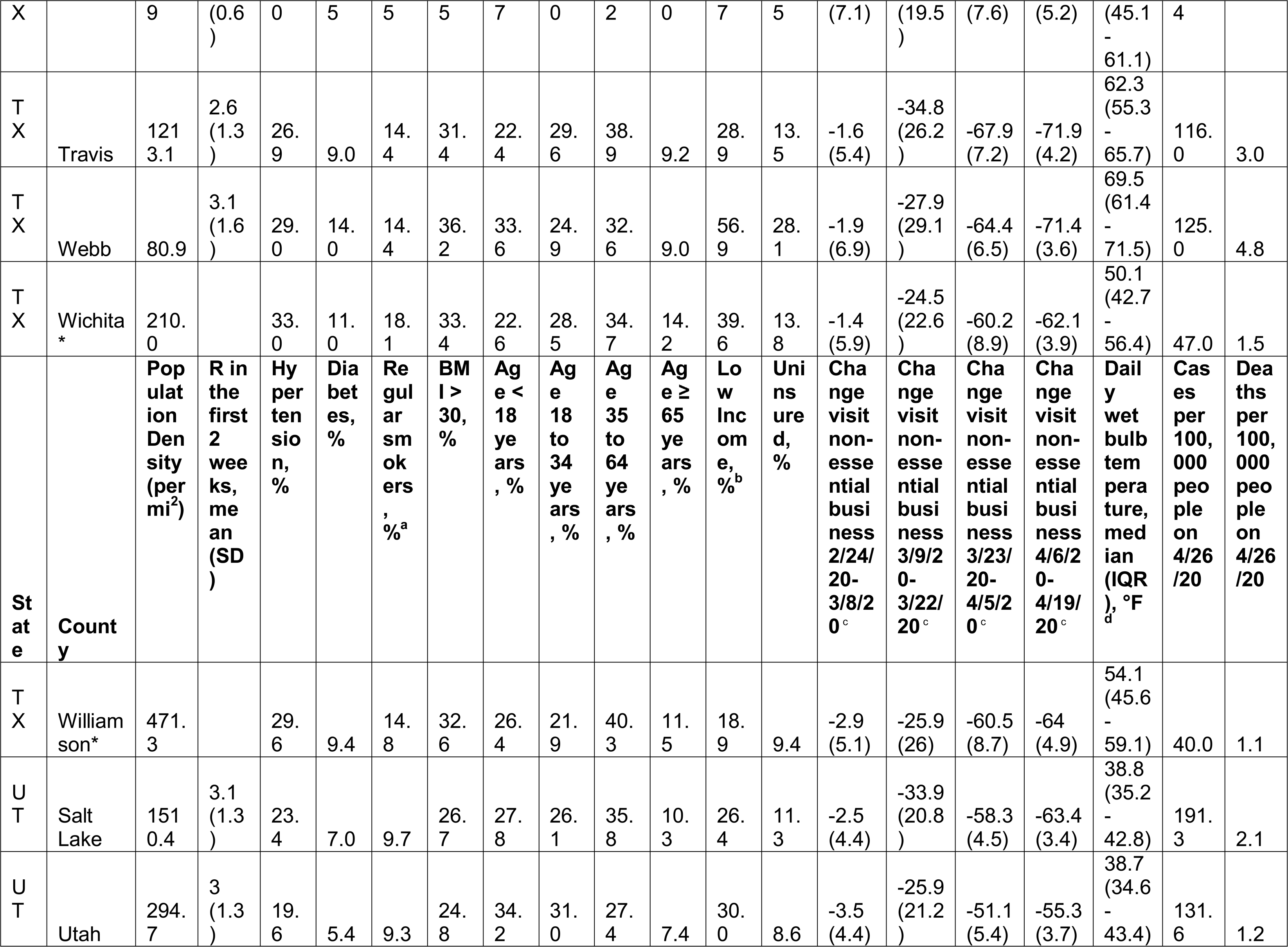
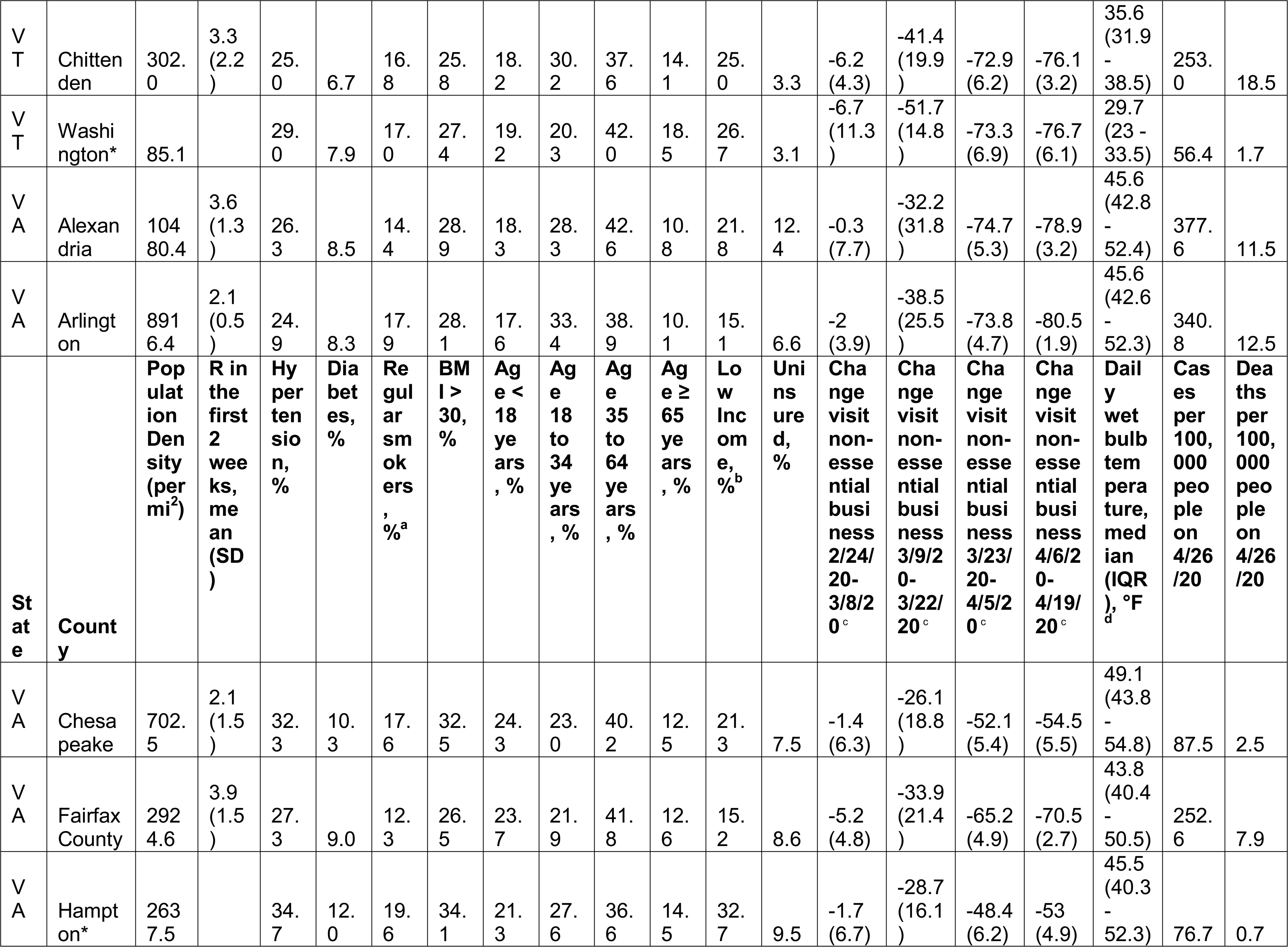
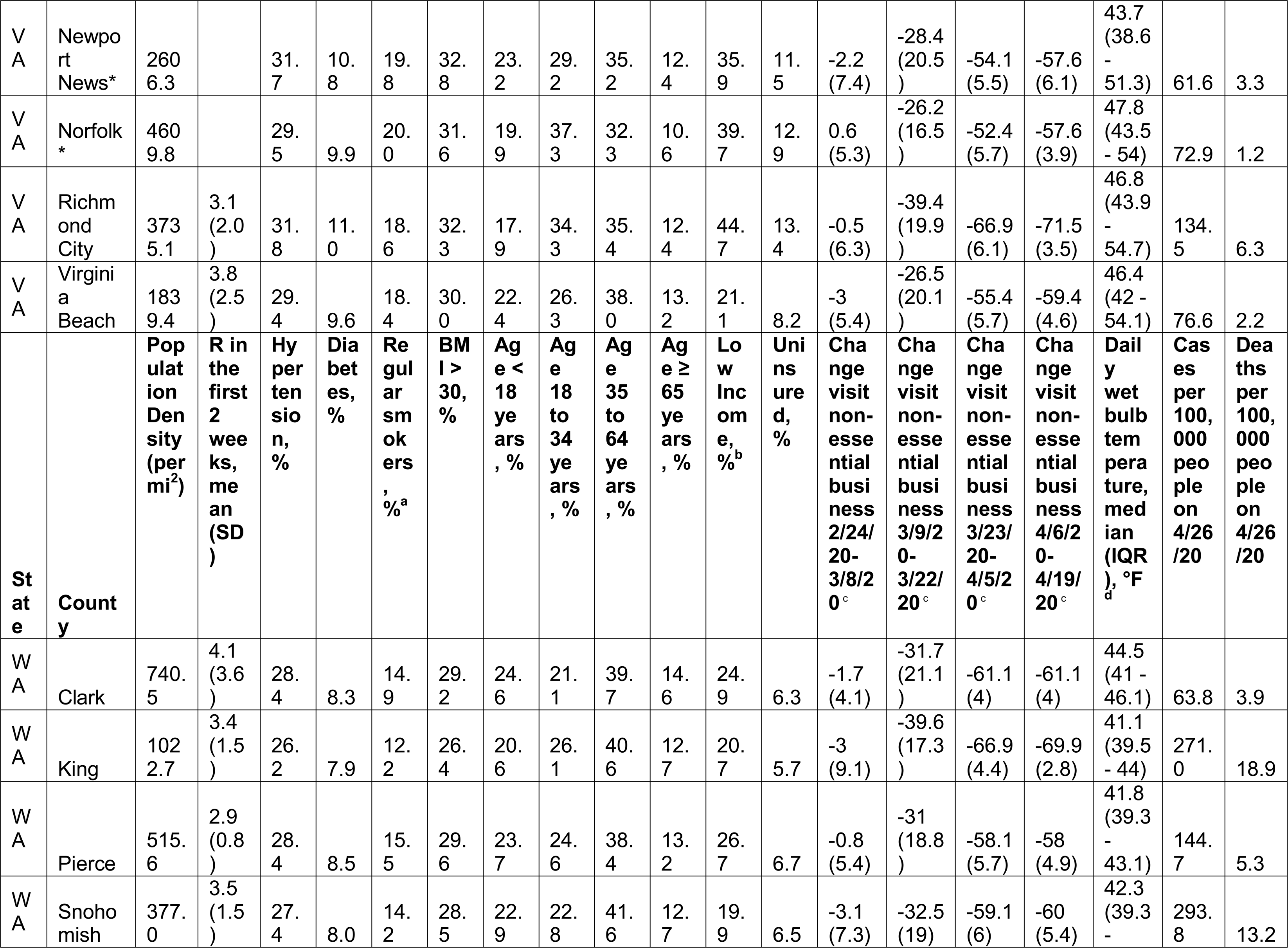
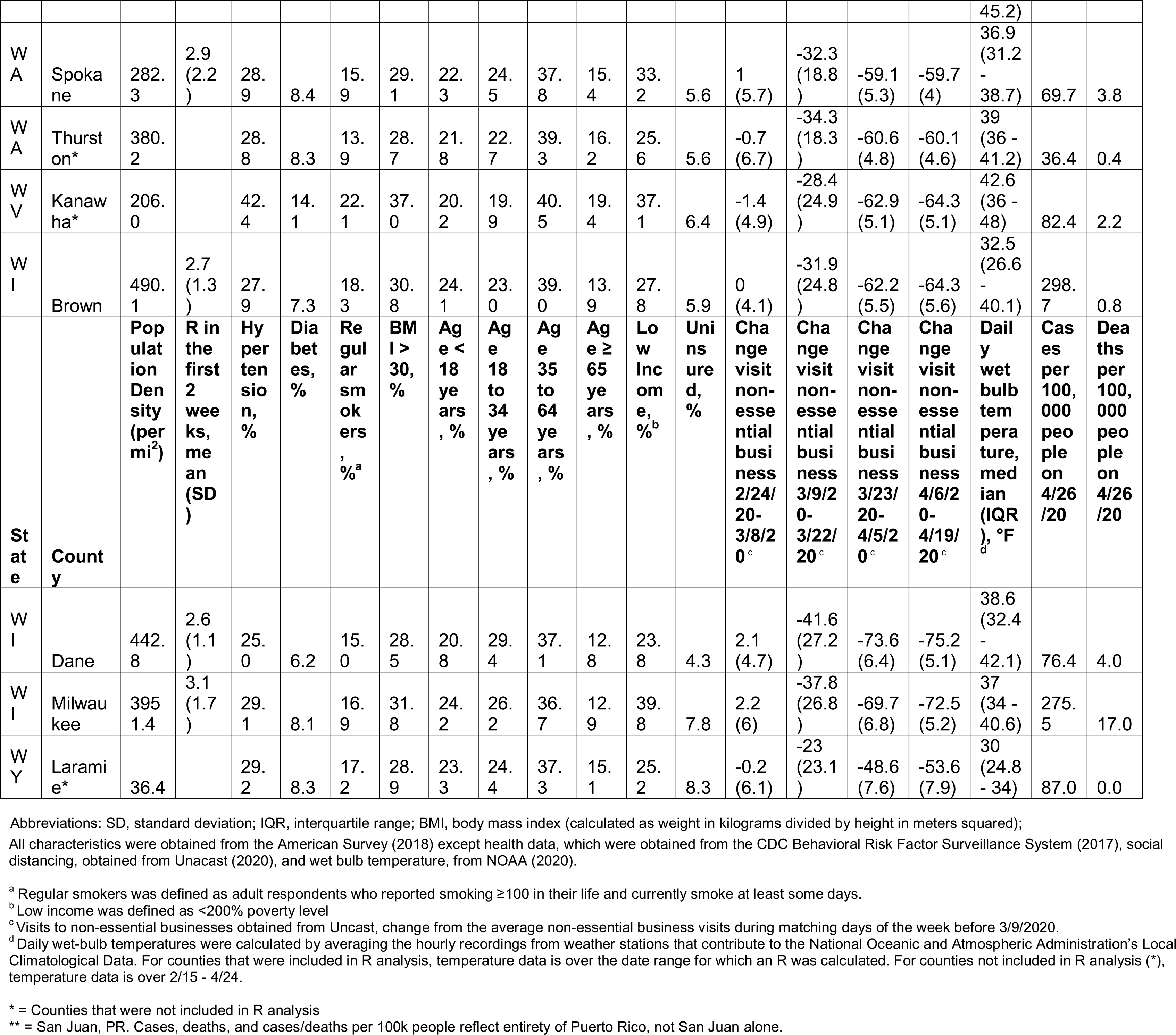
Characteristics, Demography, and Outcomes of the 260 Counties Considered in the Study.

**eFigure 1:**
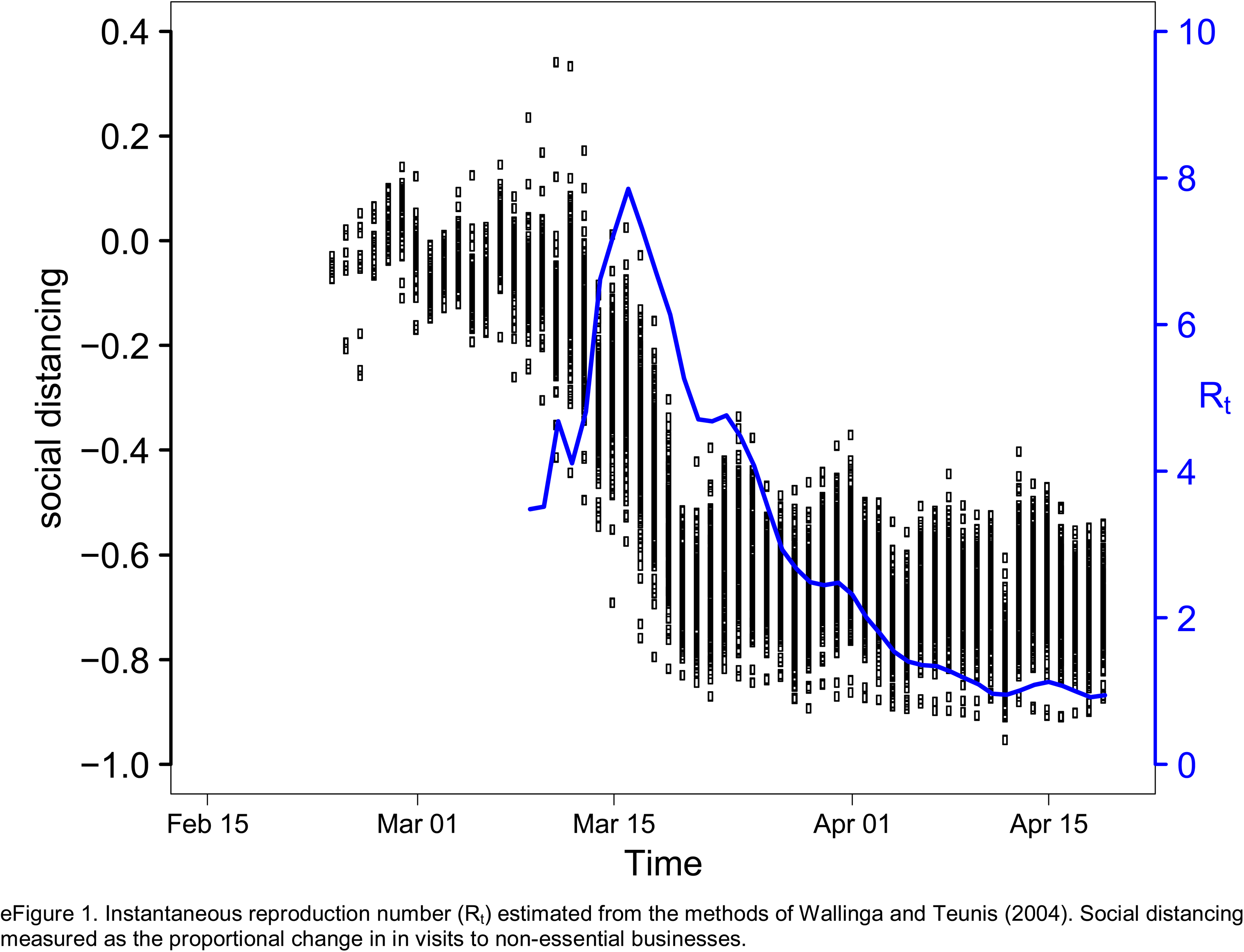
Average Instantaneous Reproduction Number (R_t_) superimposed on scatter of of social distancing over time across 211 United States counties.

**eFigure 2:**
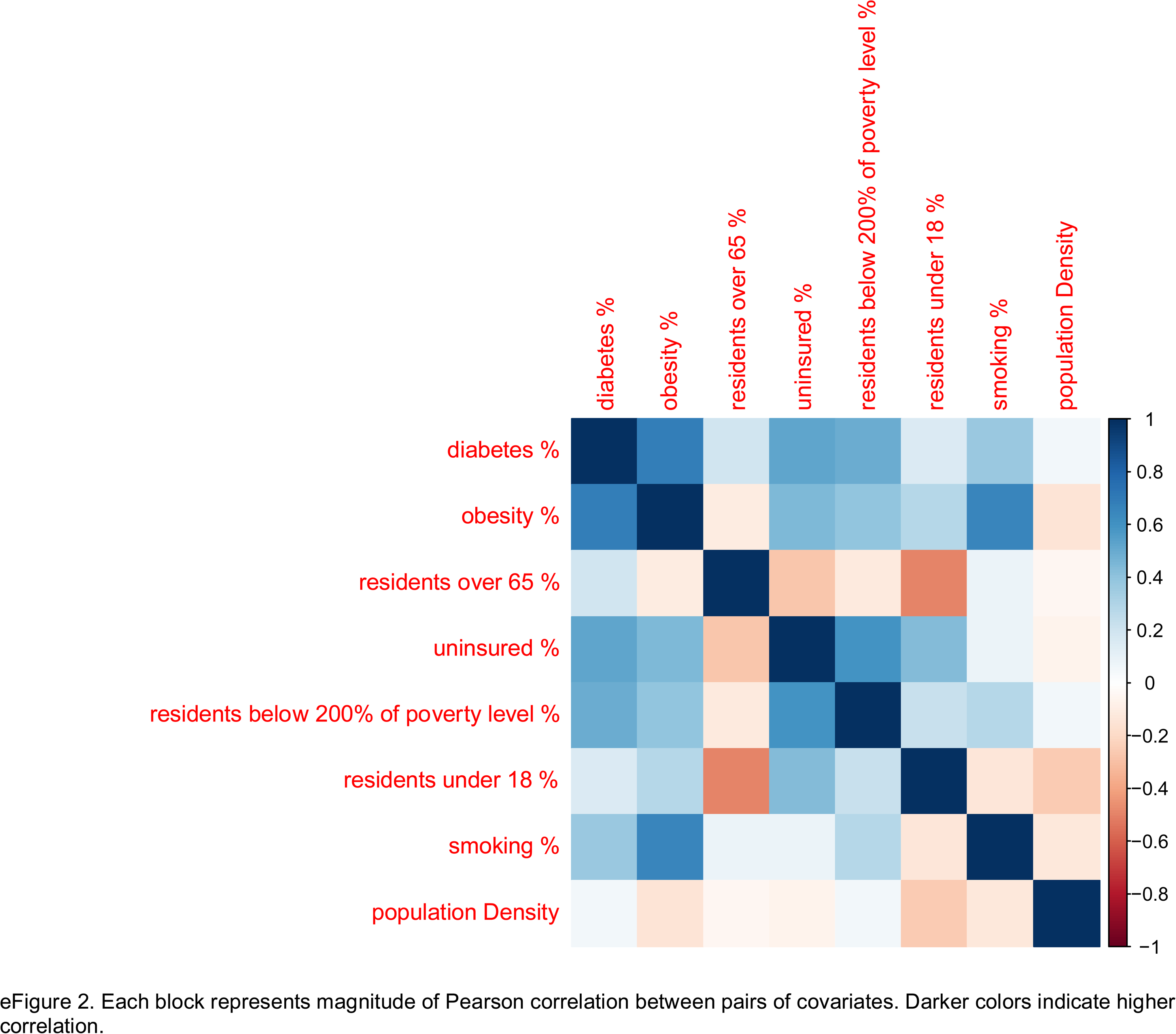
Correlation Matrix of Potential Covariates.

**eFigure 3:**
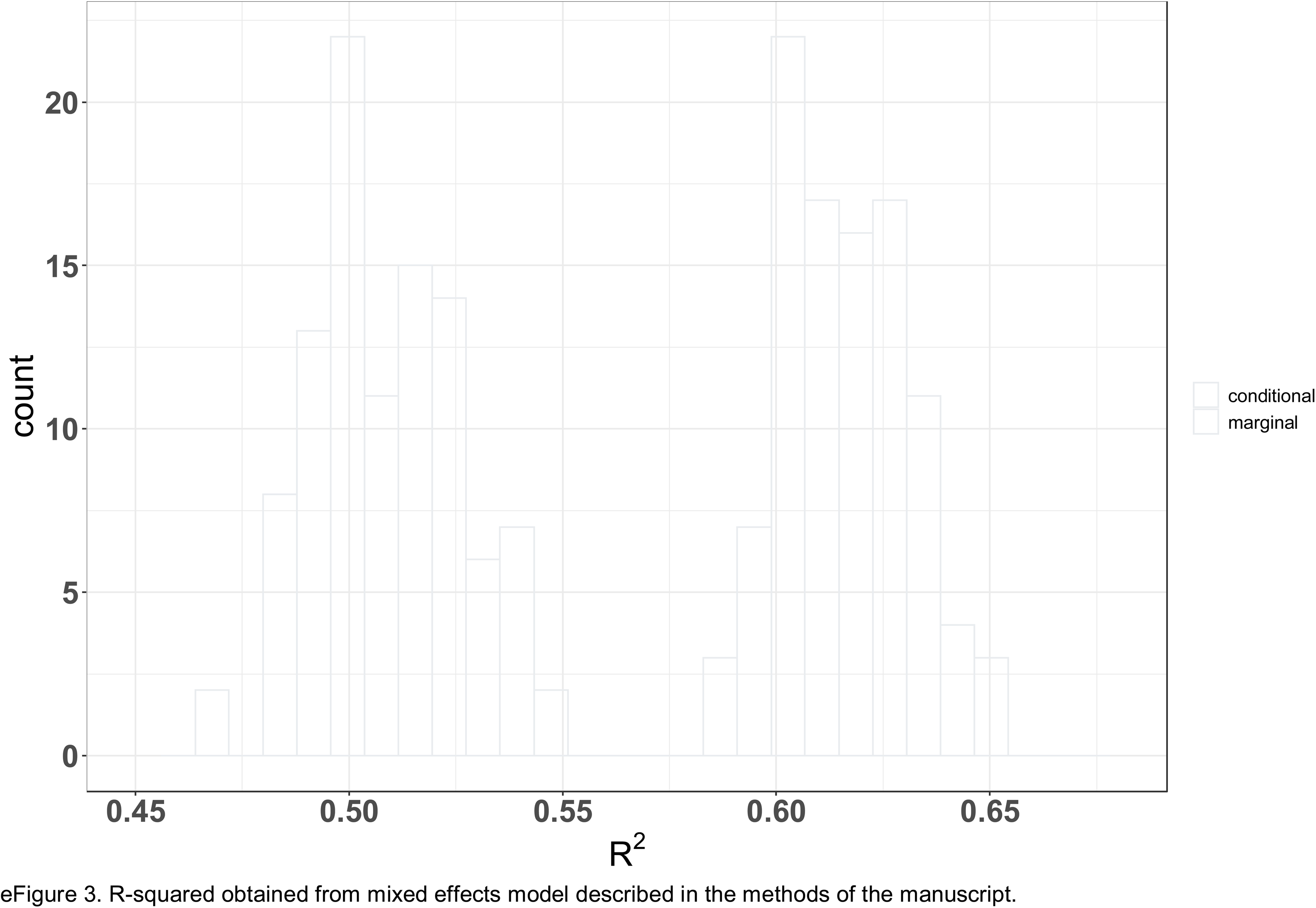
In Sample R-squared (R^2^) obtained from 100 replicates of random 70% samples of 211 United States counties.

